# Timing of First Poststroke Seizure and Risk of Seizure Recurrence by Stroke Subtype

**DOI:** 10.64898/2026.11.13.26368030

**Authors:** Kai Michael Schubert, Ingrid Berg, Mara Elena Pedrazzetti, Luisa Delazer, Erik Lindgren, Niccolò Orlandi, Stefano Meletti, Laura Abraira, Daniel Campos-Fernandez, Kazuki Fukuma, Masafumi Ihara, Vijaya Dasari, Vineet Punia, Benjamin Landau, Piergiorgio Lochner, Yaroslav Winter, Panagiota-Eleni Tsalouchidou, Yan Wang, Giovanni Merlino, Mariarosaria Valente, Carla Bentes, Matías Alet, Ana Catarina Gonçalves Franco, Ana Lúcia Oliveira, Ching Soong Khoo, Raimund Helbok, Joachim Gruber, Vasilios K. Kimiskidis, Jakob I. Doerrfuss, Vincent Leclercq, Nicolas Gaspard, Mirjam R. Heldner, Jukka Putaala, Diana Aguiar de Sousa, Jonathan M. Coutinho, María Paula Zafra-Sierra, Luis Carlos Mayor-Romero, Mark R. Keezer, Alessandro Pezzini, Rob P. W. Rouhl, John S. Duncan, Josemir W. Sander, Ronit Pressler, Eugen Trinka, Matthias J. Koepp, Barbara Tettenborn, Francesco Brigo, Marian Galovic, the SeLECT Consortium, the International Cerebral Venous Thrombosis Consortium, the MUCH-Italy Investigators, and the ILAE Acute Symptomatic Seizures Task Force

## Abstract

**Importance:** The 7-day cutoff separating acute symptomatic from unprovoked poststroke seizures guides epilepsy diagnosis and long-term antiseizure medication but is consensus based and unvalidated against recurrence risk by stroke subtype.

**Objective:** To determine whether seizure recurrence risk changes abruptly at day 7 and how it varies with first-seizure timing by stroke subtype.

**Design, Setting, and Participants:** This pooled analysis of individual participant data from 36 cohorts in 22 countries (inclusion, 1994-2025) included adults with a first seizure after ischemic stroke, intracerebral hemorrhage, aneurysmal subarachnoid hemorrhage, or cerebral venous thrombosis.

**Exposure:** Days from stroke onset (or venous thrombosis diagnosis) to first seizure.

**Main Outcomes and Measures:** The 10-year cumulative incidence of unprovoked seizure recurrence, with death as a competing event, from stabilized inverse probability of treatment– weighted Fine-Gray spline models. Risk zones were based on the likelihood of being helped vs harmed and the 60% epilepsy threshold of the International League Against Epilepsy.

**Results:** Of 2434 individuals (median [IQR] age, 63 [49-75] years; 1181 [48.5%] female), 684 (28.1%) had a seizure recurrence. Risk changed gradually with first-seizure timing, without a day-7 transition; overall 10-year risk differed by subtype (cerebral venous thrombosis, 44%; intracerebral hemorrhage, 43%; ischemic stroke, 40%; subarachnoid hemorrhage, 35%; *P* < .001). At day 7, the 10-year risk was 24% (95% CI, 17%-32%) after ischemic stroke, 30% (95% CI, 22%-38%) after intracerebral hemorrhage, 41% (95% CI, 22%-53%) after subarachnoid hemorrhage, and 58% (95% CI, 44%-71%) after cerebral venous thrombosis. The 60% threshold was crossed from day 71 after ischemic stroke, day 41 after intracerebral hemorrhage, and day 9 after cerebral venous thrombosis; after subarachnoid hemorrhage, only seizures on days 2 to 3 exceeded it. Mortality was inversely related to recurrence risk. Of 800 first seizures after day 7 following ischemic stroke or intracerebral hemorrhage, which would qualify as epilepsy under current criteria, 335 (42%) had a recurrence risk below the 60% diagnostic threshold.

**Conclusions and Relevance:** In this pooled analysis, seizure recurrence risk after stroke rose gradually, not abruptly at day 7, and differed by subtype. After external validation, subtype-specific risk zones could inform individualized decisions about long-term antiseizure medication.

**Key Points:** *Question:* Does seizure recurrence risk change abruptly at day 7, the boundary separating acute symptomatic from unprovoked seizures after stroke?

*Findings:* In this pooled analysis of individual participant data from 2434 adults with poststroke seizures, 10-year seizure recurrence risk rose gradually with first-seizure timing, without a day-7 step, and differed by stroke subtype; the 60% epilepsy threshold was reached from day 71 after ischemic stroke, day 41 after intracerebral hemorrhage, and day 9 after cerebral venous thrombosis.

*Meaning:* A single 7-day cutoff does not reflect seizure recurrence risk after stroke; subtype-specific risk zones may better guide long-term antiseizure medication decisions.

## Introduction

Stroke is the leading risk factor for epilepsy in older adults, in whom epilepsy is associated with disability, reduced quality of life, and added treatment complexity.^1-3^

The International League Against Epilepsy (ILAE) classifies seizures within 7 days of stroke as acute symptomatic and later seizures as unprovoked (remote symptomatic).^4^ The distinction has direct consequences. Epilepsy can be diagnosed after a single unprovoked seizure when the probability of further unprovoked seizures over 10 years is at least 60%, and long-term antiseizure medication (ASM) is then usually recommended.^5^ An acute symptomatic seizure carries a lower seizure recurrence risk, and long-term ASM is usually not recommended.^6^ However, the 7-day cutoff is an operational, consensus-based definition that has not been validated against seizure recurrence risk for each stroke subtype.^4,7^ Previous studies used different early-seizure definitions (7 days, 14 days, or 1 month), and subtype-specific data suggest that seizure recurrence risk changes gradually rather than on a single day.^8,9^

Dichotomizing a potentially continuous process may discard prognostic information. We therefore modeled seizure recurrence risk as a continuous function of first-seizure timing across the 4 main stroke subtypes in an international cohort (Figure 1), tested whether risk changes abruptly at day 7, and translated the resulting trajectories into clinically interpretable risk zones. The ILAE Acute Symptomatic Seizures Task Force is revising the 7-day definition, which may change when long-term ASM is started.

**Figure 1.**
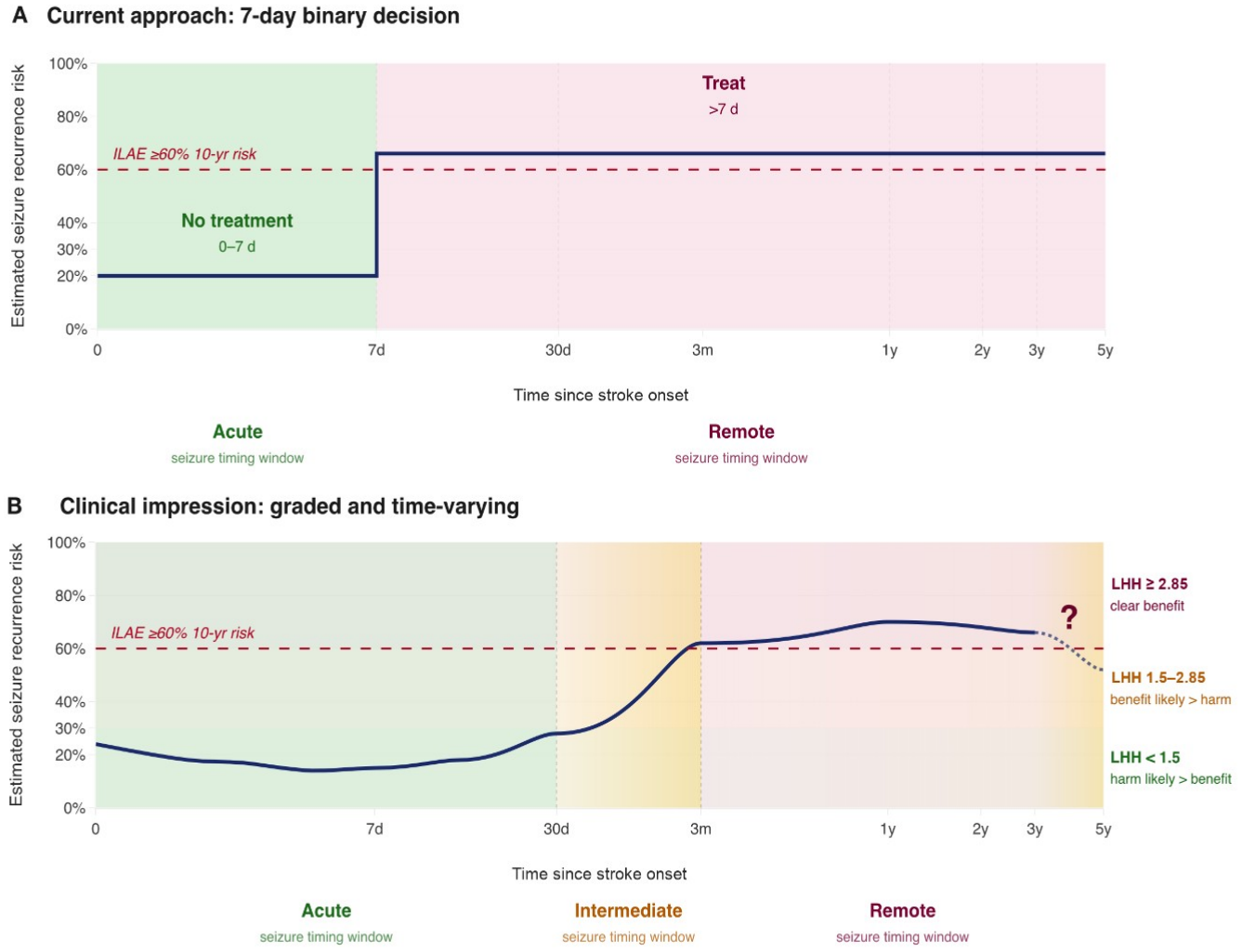
Fixed 7-Day Threshold vs Risk Trajectory of Poststroke Seizure Recurrence. A, The prevailing binary rule. Seizures within 7 days of stroke onset are classified as acute symptomatic and do not imply long-term antiseizure treatment unless additional risk features are present (imaging, electroencephalography, or a high SeLECT or CAVE score), whereas any seizure after day 7 prompts a treatment recommendation irrespective of the actual seizure recurrence risk; there is no intermediate category. B, Schematic illustration drawn from clinical impression and prior data, not fitted to the data presented here. Seizure recurrence risk is slightly elevated at day 0, remains relatively low in the acute window, rises gradually and continuously through an intermediate window (gradient shading), and reaches a broad peak in the remote window; the late long-term trajectory remains uncertain (dashed curve and question mark). The right margin illustrates the likelihood of being helped vs harmed (LHH) principle: the height of the curve at any time point indicates whether the benefit of treatment likely outweighs its harm. Under base-case assumptions, an LHH of 1.5 corresponds to a 10-year seizure recurrence risk of 31.6%, and the International League Against Epilepsy (ILAE) 60% threshold (red dashed line) corresponds to an LHH of 2.85. The fitted trajectories are shown in Figures 2 to 4.

## Methods

### Study Design and Participants

This multicenter pooled analysis included individual participant data from 36 cohorts in 22 countries on 5 continents (eTable 1 in Supplement 1). It builds on poststroke seizure prediction studies of the SeLECT Consortium, the International Cerebral Venous Thrombosis Consortium, and the Multicenter Study on Cerebral Hemorrhage in Italy (MUCH-Italy).^10-13^ Cohort inclusion periods ranged from 1994 to 2025 (eTable 2 in Supplement 1). Each cohort was approved by its local ethics committee or institutional review board, and written or verbal informed consent was obtained or waived according to local regulations (eTable 2 in Supplement 1); the pooled analysis was approved by the Cantonal Ethics Committee Zurich (BASEC 2024-02244). This study followed the Strengthening the Reporting of Observational Studies in Epidemiology (STROBE) reporting guideline.

We included adults (aged ≥18 years) with a first-ever acute ischemic stroke, spontaneous intracerebral hemorrhage (ICH), aneurysmal subarachnoid hemorrhage (SAH), or cerebral venous thrombosis (CVT), hereafter the main stroke subtypes, and at least 1 clinically confirmed seizure. We excluded individuals with preexisting epilepsy, seizures unrelated to the stroke, or follow-up shorter than 30 days after the index stroke (eFigure 1 in Supplement 1). Sex was recorded by the contributing centers from medical records. Data on race and ethnicity were not collected consistently across the international cohorts and were therefore not analyzed.

### Exposure and Outcomes

The exposure was first-seizure timing: the interval in days between stroke onset (date of diagnosis for CVT; last-seen-well date if onset was unclear) and the first documented seizure. For CVT, seizures before the diagnosis date were assigned to day 0. Acute symptomatic and unprovoked seizures were defined according to ILAE definitions.^4,5^ The primary outcome was seizure recurrence, defined as an unprovoked seizure according to ILAE criteria and ascertained according to the protocol of each cohort (eMethods in Supplement 1). It was summarized as the 10-year cumulative incidence after the first seizure, with all-cause death as the competing event. The secondary outcome was all-cause mortality.

### Risk Zones

Because seizure recurrence risk may rise continuously rather than stepwise (Figure 1), we defined risk zones with the likelihood of being helped vs harmed (LHH) by long-term ASM, a transparent and updatable framework,^14^ recently discussed for epilepsy.^15^ Its base-case assumptions, a relative risk reduction of 0.25 and a number needed to harm of 19, derive from randomized ASM trial data^16,17^ (eMethods in Supplement 1); alternative values are shown in eTable 3 in Supplement 1. The lower boundary, an LHH of 1.5 (10-year cumulative incidence, 31.6%), is the point at which benefit first exceeds harm by 3:2. The upper boundary is the ILAE threshold for diagnosing epilepsy after a single unprovoked seizure (≥60%),^5^ which corresponds to an LHH of 2.85. First seizures were classified as carrying low (<31.6%), moderate (31.6% to <60%), or high (≥60%) seizure recurrence risk. For each subtype, the days on which the spline-smoothed recurrence curve crossed these thresholds defined the zone boundaries.

### Statistical Analysis

We used Fine-Gray subdistribution hazard models,^18^ with time zero at the first poststroke seizure, because the clinically relevant quantity is the absolute seizure recurrence probability under competing mortality. Because ASM exposure was not randomized, we applied stabilized inverse probability of treatment weighting (covariate balance in eTable 4 in Supplement 1).^19^ Stabilized weights were used for all subtypes because ASM treatment was nearly universal after SAH (95%), for which unstabilized weights reduced the effective sample size to 12%; stabilization restored it to 99% (eMethods in Supplement 1). First-seizure timing was modeled with natural cubic splines on log(1 + day), with 3 degrees of freedom (5 for SAH, to accommodate its early and late peaks), within the same weighted model for all subtypes. The 95% CIs of the curves and zone boundaries were obtained by refitting the model in 1000 bootstrap resamples.

To compare the 7-day boundary with data-driven alternatives, we ranked binary cutoffs from day 1 to day 90 by the bayesian information criterion (BIC; eTable 5 in Supplement 1). This analysis was restricted to ischemic stroke and ICH because the SAH and CVT samples were smaller. Prespecified analyses tested whether the 10-year cumulative incidence differed across risk zones within each subtype (Gray K-sample test) and whether seizure recurrence risk and its trajectory differed between subtypes independently of timing (4-sample Gray test, timing-adjusted Fine-Gray Wald test, spline interaction, and pseudo-value generalized additive model; eTable 6 in Supplement 1). Zone stability was assessed with 1000-iteration bootstraps of the boundary days and of the fixed classification (eTable 7 and eFigures 3 and 4 in Supplement 1). A leave-one-cohort-out analysis was restricted to cohorts contributing at least 25 individuals to a subtype (29 of 51 subtype-cohort units; 2184 of 2434 individuals [90%]) (eTables 1 and 8 in Supplement 1).

Missing covariate values were imputed with the modal or median value (eMethods in Supplement 1). Statistical tests were 2-sided. Analyses used R version 4.5.2 (R Foundation for Statistical Computing).

## Results

### Study Population

Of 2978 individuals with a first poststroke seizure, 2434 met the eligibility criteria (ischemic stroke, 931; ICH, 715; SAH, 338; CVT, 450) (Table 1; eFigure 1 and eTables 1 and 9 in Supplement 1). The median (IQR) age was 63 (49-75) years, and 1253 (51.5%) were male. The first seizure occurred within 7 days in 467 individuals (50.2%) with ischemic stroke, 379 (53.0%) with ICH, 234 (69.2%) with SAH, and 414 (92.0%) with CVT (Figure 2A and B; Figure 3A and B). The median (IQR) follow-up after the first seizure was 1.0 (0.3-3.4) years. During follow-up, 684 individuals (28.1%) had a seizure recurrence and 311 (12.8%) died without a prior recurrence.

**Table 1.** Characteristics of the Study Population by Stroke Subtype.

| Characteristic | Ischemic stroke (n = 931) | ICH (n = 715) | SAH (n = 338) | CVT (n = 450) |
|---|---|---|---|---|
| **Study population** |  |  |  |  |
| Records screened, No. | 1106 | 956 | 390 | 526 |
| Excluded, No. | 175 | 241 | 52 | 76 |
| Missing follow-up time | 88 | 219 | 9 | 3 |
| Missing first-seizure timing | 69 | 3 | 2 | 5 |
| Postseizure follow-up <30 d (day 0-30 window) | 18 | 19 | 41 | 68 |
| Eligible for analysis, No. | 931 | 715 | 338 | 450 |
| Contributing cohorts (centers), No. | 17 | 10 | 9 | 15 |
| **Demographics** |  |  |  |  |
| Age, median (IQR), y | 70.0 (58.0-79.0) | 68.0 (56.0-78.0) | 54.1 (46.3-65.0) | 41.0 (30.0-53.0) |
| Male sex | 583 (62.6) | 413 (57.8) | 117 (34.6) | 140 (31.1) |
| **Clinical characteristics** |  |  |  |  |
| *Ischemic stroke* |  |  |  |  |
| NIHSS score 4-10 (moderate) | 305 (32.8) | NA | NA | NA |
| NIHSS score >10 (severe) | 389 (41.8) | NA | NA | NA |
| Large-artery atherosclerosis | 227 (24.4) | NA | NA | NA |
| Cardioembolic mechanism | 282 (30.3) | NA | NA | NA |
| Small-vessel disease | 62 (6.7) | NA | NA | NA |
| MCA territory involvement | 719 (77.2) | NA | NA | NA |
| Cortical involvement | 716 (76.9) | NA | NA | NA |
| Age ≥65 y | 561 (60.3) | NA | NA | NA |
| *Intracerebral hemorrhage* |  |  |  |  |
| NIHSS score 4-10 (moderate) | NA | 182 (25.5) | NA | NA |
| NIHSS score >10 (severe) | NA | 330 (46.2) | NA | NA |
| Lobar hemorrhage | NA | 437 (61.1) | NA | NA |
| Hematoma volume, median (IQR), mL | NA | 25.3 (8.6-50.0) | NA | NA |
| Neurosurgical intervention | NA | 110 (15.4) | NA | NA |
| *Subarachnoid hemorrhage* |  |  |  |  |
| Modified Rankin Scale score at discharge, median (IQR) | NA | NA | 3 (1-5) | NA |
| VASOGRADE score, median (IQR) | NA | NA | 1 (1-2) | NA |
| Aneurysm treatment or surgery | NA | NA | 313 (92.6) | NA |
| *Cerebral venous thrombosis* |  |  |  |  |
| ICH at presentation | NA | NA | NA | 239 (53.1) |
| Subdural hematoma | NA | NA | NA | 28 (6.2) |
| **Treatment** |  |  |  |  |
| Any antiseizure medication | 797 (85.6) | 607 (84.9) | 321 (95.0) | 388 (86.2) |
| Reperfusion treatment (ischemic stroke only) | 357 (38.3) | NA | NA | NA |
| Decompressive surgery (CVT only) | NA | NA | NA | 30 (6.7) |
| **First-seizure timing** |  |  |  |  |
| First-seizure day, median (IQR) | 7 (0-280) | 5 (1-193) | 1 (0-25) | 0 (0-0) |
| First seizures on days 0-7, median (IQR) | 0.0 (0.0-2.0) | 1.0 (0.0-2.0) | 0.0 (0.0-1.0) | 0.0 (0.0-0.0) |
| First seizures after day 7, median (IQR) | 280.5 (124-586) | 214 (90-540) | 184.5 (58-487) | 256.0 (75-739) |
| First seizure on days 0-7 (acute symptomatic) | 467 (50.2) | 379 (53.0) | 234 (69.2) | 414 (92.0) |
| First seizure on day ≥8 | 464 (49.8) | 336 (47.0) | 104 (30.8) | 36 (8.0) |
| **Missing data** |  |  |  |  |
| Age | 0 (0.0) | 0 (0.0) | 0 (0.0) | 8 (1.8) |
| Sex | 0 (0.0) | 0 (0.0) | 0 (0.0) | 0 (0.0) |
| Stroke severity (NIHSS score) | 0 (0.0) | 5 (0.7) | NA | NA |
| Stroke mechanism (ischemic stroke) | 1 (0.1) | NA | NA | NA |
| Hematoma volume (ICH) | NA | 159 (22.2) | NA | NA |
| Modified Rankin Scale score at discharge (SAH) | NA | NA | 95 (28.1) | NA |
| VASOGRADE score (SAH) | NA | NA | 35 (10.4) | NA |
| Any antiseizure medication | 0 (0.0) | 113 (15.8) | 0 (0.0) | 0 (0.0) |
| **Outcomes and follow-up** |  |  |  |  |
| Follow-up, median (IQR), y | 1.0 (0.3-3.6) | 0.5 (0.1-2.2) | 3.3 (0.9-6.1) | 1.0 (0.5-2.2) |
| Seizure recurrence (primary outcome) | 277 (29.8) | 209 (29.2) | 101 (29.9) | 97 (21.6) |
| Death without prior seizure recurrence | 90 (9.7) | 159 (22.2) | 34 (10.1) | 28 (6.2) |

Abbreviations: CVT, cerebral venous thrombosis; ICH, intracerebral hemorrhage; MCA, middle cerebral artery; NA, not applicable; NIHSS, National Institutes of Health Stroke Scale; SAH, subarachnoid hemorrhage.
Values are No. (%) unless otherwise indicated. Follow-up is measured from the first poststroke seizure. Four individuals whose recorded follow-up was entered as a negative value were confirmed by the contributing centers to have 1 day of follow-up and are analyzed as such. Antiseizure medication denotes any antiseizure medication recorded by the contributing cohort, started either acutely after the stroke or after a later seizure. Modified Rankin Scale and VASOGRADE are ordinal scales; medians are shown as whole numbers where every quantile is an integer. Exclusion reasons are hierarchical and mutually exclusive and sum to the number excluded (eTable 9 in Supplement 1); missing data rows are descriptive, with percentages of the number eligible for analysis.

**Figure 2.**
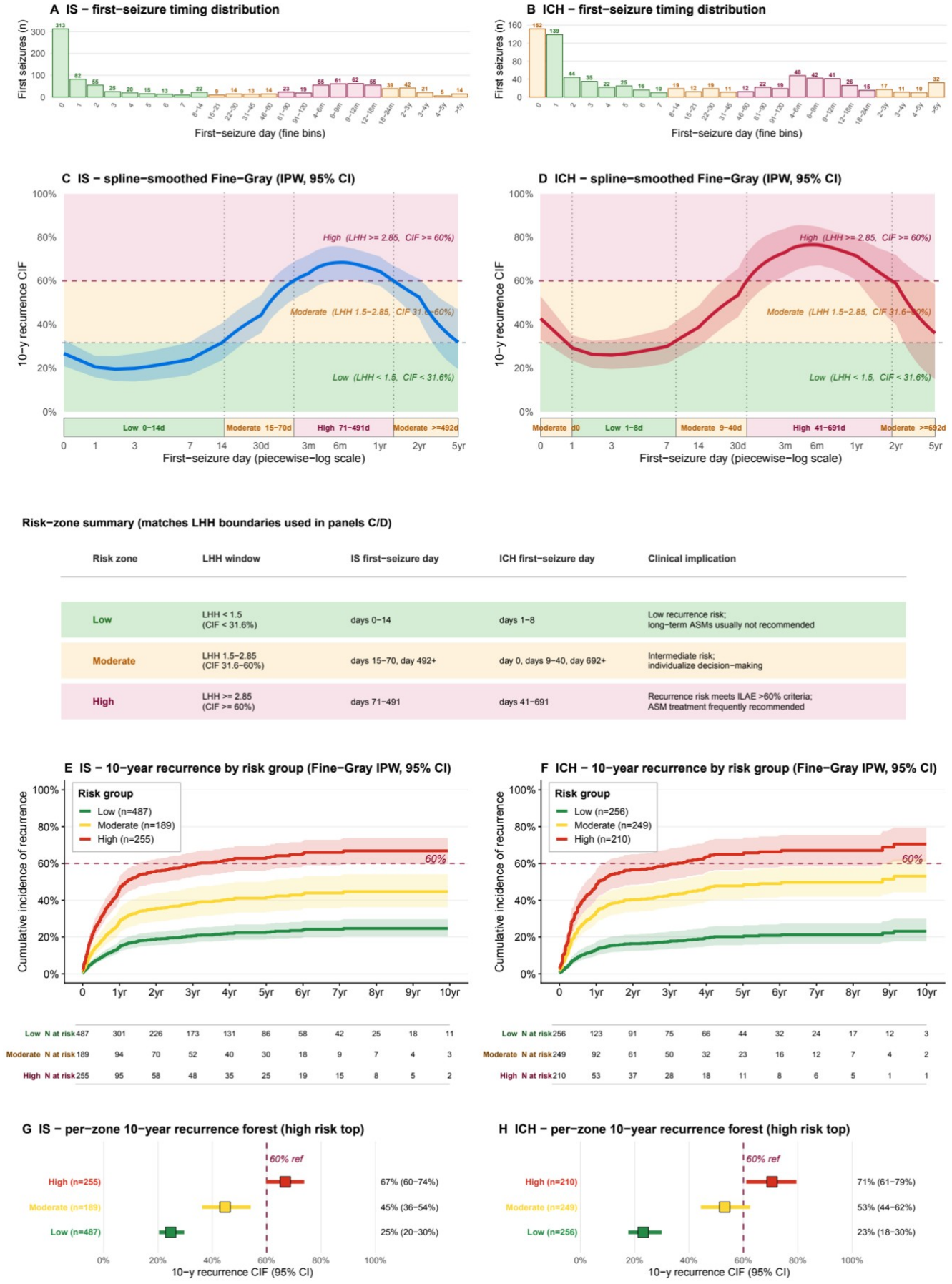
Seizure Recurrence by Timing of the First Seizure After Ischemic Stroke and Intracerebral Hemorrhage. Data are shown for ischemic stroke (IS; n = 931) and intracerebral hemorrhage (ICH; n = 715). A and B, Distribution of first-seizure timing, with bars colored by risk zone. C and D, Stabilized inverse probability–weighted (IPW) Fine-Gray spline estimate of the 10-year cumulative incidence function (CIF) of seizure recurrence by first-seizure day (solid line; shading, 95% bootstrap interval from 1000 resamples), shown against risk bands based on the likelihood of being helped vs harmed (LHH): green, low risk (CIF <31.6%; LHH <1.5); yellow, moderate risk (CIF 31.6% to <60%; LHH 1.5-2.85); red, high risk (CIF ≥60%; LHH ≥2.85). Vertical dotted lines and the strip below each panel mark the resulting zones (IS: low, days 0-14; moderate, days 15-70 and ≥492; high, days 71-491. ICH: moderate, day 0, days 9-40, and ≥692; low, days 1-8; high, days 41-691). The table between the upper and lower rows of panels restates the LHH thresholds and the resulting zone days for both subtypes. E and F, Ten-year cumulative incidence of seizure recurrence by risk zone (Fine-Gray IPW step curves), with numbers at risk from 0 to 10 years. G and H, Ten-year cumulative incidence of seizure recurrence by risk zone (point estimates with 95% CIs; high-risk zone at top; dashed line, 60% International League Against Epilepsy [ILAE] threshold). Death without prior seizure recurrence was the competing event.

**Figure 3.**
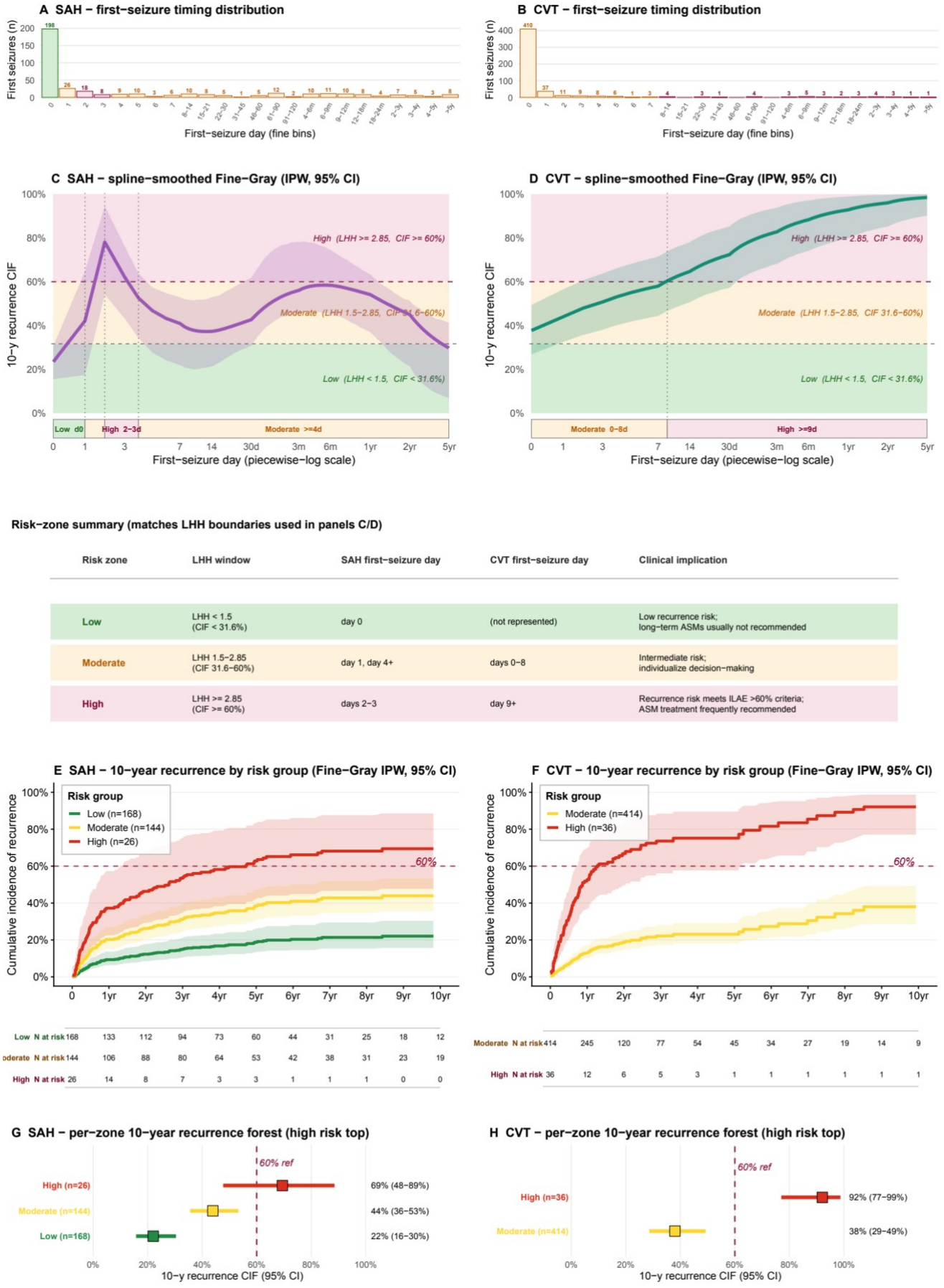
Seizure Recurrence by Timing of the First Seizure After Subarachnoid Hemorrhage and Cerebral Venous Thrombosis. Data are shown for subarachnoid hemorrhage (SAH; n = 338) and cerebral venous thrombosis (CVT; n = 450), using the same layout as Figure 2. A and B, Distribution of first-seizure timing, with bars colored by risk zone. C and D, Stabilized inverse probability–weighted (IPW) Fine-Gray spline estimate of the 10-year cumulative incidence function (CIF) of seizure recurrence (solid line; shading, 95% bootstrap interval from 1000 resamples) against risk bands based on the likelihood of being helped vs harmed (LHH; green, low; yellow, moderate; red, high) and the resulting zones (SAH: low, day 0; moderate, day 1 and days ≥4; high, days 2-3. CVT: moderate, days 0-8; high, from day 9). The table between the upper and lower rows of panels restates the LHH thresholds and the resulting zone days for SAH and CVT. E and F, Ten-year cumulative incidence of seizure recurrence by risk zone (Fine-Gray IPW step curves), with numbers at risk from 0 to 10 years. G and H, Ten-year cumulative incidence by risk zone (point estimates with 95% CIs; dashed line, 60% International League Against Epilepsy threshold). Estimates for the SAH high-risk zone (days 2-3) and for late first seizures after CVT rest on small numbers and warrant caution. Death without prior seizure recurrence was the competing event.

### First-Seizure Timing and Seizure Recurrence Risk

After ischemic stroke and ICH, seizure recurrence risk was low for very early first seizures and then rose steadily (Figure 2C and D). The estimated 10-year seizure recurrence risk peaked at 68% (95% CI, 60%-76%) for first seizures at approximately 6 months (day 188) after ischemic stroke and at 77% (95% CI, 67%-85%) at approximately 5 to 6 months (day 167) after ICH, and declined gradually for later first seizures. Unlike after ischemic stroke, first seizures on the day of ICH were associated with a higher seizure recurrence risk (43%; 95% CI, 33%-53%) than those on days 1 to 7 (eg, 26%; 95% CI, 19%-33% at the day 3 nadir).

Because most first seizures after SAH and CVT occurred within 7 days, later estimates were less precise. After SAH, seizure recurrence risk peaked early on days 2 to 3 (78%; 95% CI, 54%-95% on day 2), declined, and rose again later without reaching 60% (Figure 3C). After CVT, it increased steadily from moderate risk for first seizures on days 0 to 8 to high risk from day 9 (Figure 3D).

Pooled across first-seizure timings, the 10-year seizure recurrence risk was highest after CVT (44%; 95% CI, 34%-55%), followed by ICH (43%; 95% CI, 38%-48%), ischemic stroke (40%; 95% CI, 36%-44%), and SAH (35%; 95% CI, 29%-42%) (Figure 4). The absolute seizure recurrence risk and its trajectory differed between subtypes (Gray test, *P* < .001; timing-adjusted Fine-Gray model, *P* = .003; trajectory interaction, *P* < .001; pseudo-value trajectory model, *P* = .004; eTable 6 in Supplement 1).

**Figure 4.**
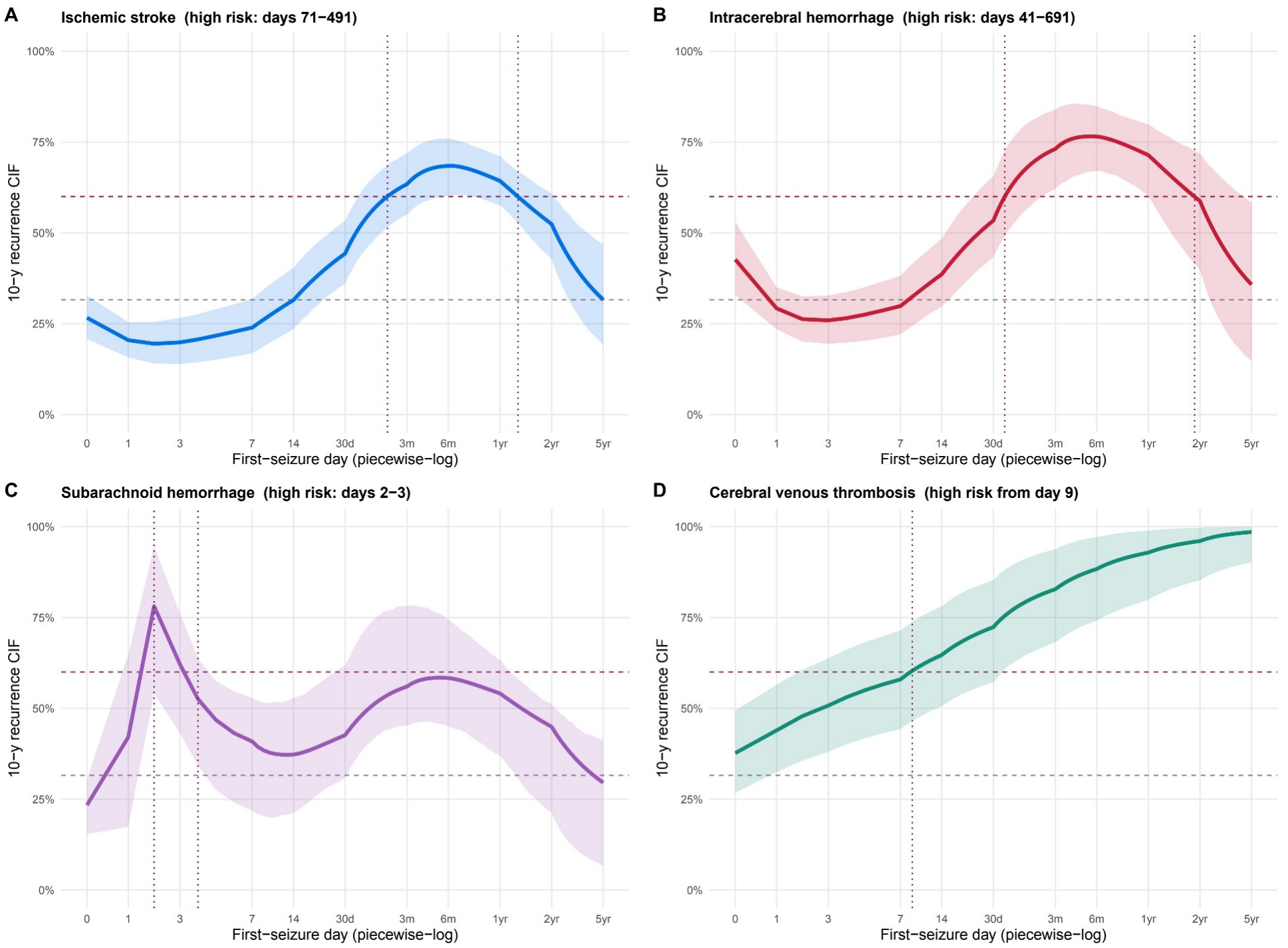
Seizure Recurrence Risk Trajectory by Stroke Subtype. For each stroke subtype, the 10-year cumulative incidence function (CIF) of seizure recurrence is shown as a function of first-seizure timing (piecewise logarithmic axis). Solid lines show the stabilized inverse probability–weighted (IPW) Fine-Gray spline point estimate (identical to Figure 2C and D and Figure 3C and D); shading shows the 95% percentile interval of the same model refitted in 1000 bootstrap resamples (window stratified for the day 0–heavy subarachnoid hemorrhage [SAH] and cerebral venous thrombosis [CVT] cohorts). Horizontal dashed lines mark the 31.6% (likelihood of being helped vs harmed [LHH], 1.5) and 60% (International League Against Epilepsy; high risk) thresholds; vertical dotted lines mark the days on which the curve enters and leaves the high-risk zone (ischemic stroke, days 71-491; intracerebral hemorrhage [ICH], days 41-691; SAH, days 2-3; CVT, from day 9). Death without prior seizure recurrence was the competing event.

### The 7-Day Cutoff and Risk Zones

Seizure recurrence curves after ischemic stroke and ICH changed smoothly, without an abrupt transition (Figure 2C and D). At day 7, the 10-year seizure recurrence risk was 24% (95% CI, 17%-32%) after ischemic stroke and 30% (95% CI, 22%-38%) after ICH, both below the 60% threshold. The risk first reached 60% at day 71 after ischemic stroke (bootstrap 95% interval, day 43-138) and at day 41 after ICH (bootstrap 95% interval, day 24-77). The BIC-preferred binary cutoff was day 20 for ischemic stroke (ΔBIC vs day 7, −10.7) and day 12 for ICH (ΔBIC, −9.6) (eTable 5 in Supplement 1). Bootstrap medians of the zone entry days were within 1 day of the point estimates for all 4 subtypes (eFigure 3 and eTable 7 in Supplement 1).

After ischemic stroke, seizure recurrence risk was low for first seizures on days 0 to 14, moderate on days 15 to 70, and high on days 71 to 491; from day 492 (approximately 16 months), it fell back to moderate (Figure 2C). After ICH, risk was moderate on day 0, low on days 1 to 8, moderate on days 9 to 40, and high on days 41 to 691, reverting to moderate from day 692 (approximately 23 months) (Figure 2D). After SAH, risk was high only on days 2 to 3, and after CVT, from day 9 onward (Figure 3C and D).

The 10-year cumulative incidence of seizure recurrence differed across risk zones in each subtype (*P* < .001 for all; Figure 2E-H and Figure 3E-H; unadjusted Aalen-Johansen estimates in eFigure 2 in Supplement 1). After ischemic stroke, it was 25% (95% CI, 20%-30%) in the low-risk, 45% (95% CI, 36%-54%) in the moderate-risk, and 67% (95% CI, 60%-74%) in the high-risk zone; after ICH, 23% (95% CI, 18%-30%), 53% (95% CI, 44%-62%), and 71% (95% CI, 61%-79%); and after SAH, 22% (95% CI, 16%-30%), 44% (95% CI, 36%-53%), and 69% (95% CI, 48%-89%). After CVT, it was 38% (95% CI, 29%-49%) in the moderate-risk and 92% (95% CI, 77%-99%) in the high-risk zone. The zones separated significantly in 99.9% to 100% of bootstrap resamples, were monotonically ordered in 96% (SAH) to 100% of resamples (eFigure 4 and eTable 7 in Supplement 1), and differed significantly in every leave-one-cohort-out analysis (eTable 8 in Supplement 1).

Of 800 first seizures after day 7 following ischemic stroke or ICH, which the current rule classifies as unprovoked, 335 (42%) had an estimated 10-year seizure recurrence risk below 60% (209 of 464 [45%] after ischemic stroke; 126 of 336 [38%] after ICH) (eTable 10 in Supplement 1). Zone-specific cumulative incidence curves can be explored in an interactive web tool (https://predictepilepsy.com/calc-post-stroke-recurrence/).

### Mortality

All-cause mortality broadly showed the opposite pattern (eFigures 5 and 6 in Supplement 1). After ischemic stroke, 10-year mortality was 27% (95% CI, 19%-38%) in the low-risk, 21% (95% CI, 12%-35%) in the moderate-risk, and 14% (95% CI, 8%-25%) in the high-risk zone. After ICH, it was 64% (95% CI, 56%-72%) for first seizures on days 1 to 8 (low risk), 16% (95% CI, 11%-25%) in the moderate-risk, and 17% (95% CI, 10%-26%) in the high-risk zone. After SAH, it was 16% (95% CI, 10%-25%), 12% (95% CI, 7%-20%), and 6% (95% CI, 1%-35%), respectively. After CVT, mortality was 24% (95% CI, 15%-38%) in the moderate-risk zone, with no deaths in the high-risk zone.

## Discussion

In this international pooled analysis, seizure recurrence risk after a first poststroke seizure did not change abruptly at day 7. It rose gradually with first-seizure timing, followed a distinct trajectory for each stroke subtype, and, after ischemic stroke and ICH, crossed the 60% epilepsy threshold only weeks after day 7, at subtype-specific times. Consequently, 2 in 5 first seizures after day 7 in these subtypes, which would currently be diagnosed as epilepsy, carried a seizure recurrence risk below this threshold.

Early definitions of acute symptomatic seizures ranged from 24 hours to 1 month after the insult.^20,21^ The ILAE adopted 7 days for consistency of classification rather than as a threshold validated against seizure recurrence for each stroke subtype.^4^ That choice did not rest on robust evidence^22^ but on a then-prevailing cutoff.^6,23,24^ An influential report using this rule found lower seizure recurrence but higher mortality after acute symptomatic seizures but was small, pooled different causes, and predated modern stroke care.^6^

The gradual rise in risk is consistent with 2 overlapping processes. Early after stroke, cell death, release of excitatory neurotransmitters, inflammation, and blood-brain barrier breakdown can directly provoke seizures, which carry a relatively low seizure recurrence risk once the acute phase resolves.^25,26^ Epileptogenesis, by contrast, unfolds over weeks to years.^25^ Because these processes overlap, no single day marks a biologically meaningful transition, and even the statistically best binary cutoffs (day 20 after ischemic stroke, day 12 after ICH) would reduce a continuous gradient to an artificial step. Subtype differences are plausible given differences in mechanism, lesion location and size, and the balance of ischemic and hemorrhagic injury. The higher risk after CVT and ICH than after ischemic stroke is well recognized,^13,27-29^ and the lower risk after SAH aligns with the absence of large intracerebral lesions in many patients.^30^

Rather than replace one cutoff with another, we mapped the continuous curves into risk zones linked to treatment decisions.^14,17,31^ Low-risk seizures are unlikely to benefit consistently from long-term ASM. High-risk seizures meet the ILAE threshold for diagnosing epilepsy after a single seizure, at which most clinicians would offer treatment. In the moderate zone, net benefit is uncertain and decisions should be individualized and shared. People for whom a further seizure would have serious consequences, such as injury, loss of driving privileges, occupational consequences, or high anxiety, may accept treatment at lower risk, whereas those concerned about adverse effects, polypharmacy, or pregnancy may require higher expected benefit; frailty can argue either way.^32,33^ Seizure-risk tolerance varies widely, and graphical risk communication is welcomed by patients.^34^ The framework imposes no new diagnostic category, its assumptions are explicit and can be updated as trial data emerge,^35,36^ and the interactive tool lets users adjust them. If a dichotomy is retained for simplicity, it could be subtype specific, for example the day on which the 10-year seizure recurrence risk first exceeds 60%.

Under the current rule, these lower-risk late seizures lead to an epilepsy diagnosis and often long-term ASM. Such potential overdiagnosis and overtreatment can expose patients to stigma, adverse effects, driving and occupational restrictions, and health care costs, with limited expected benefit. Whether a risk-based approach reduces this burden without increasing seizure-related harm should be evaluated prospectively.

Seizure recurrence risk appeared to fall again when the first seizure occurred after approximately 16 months (ischemic stroke) or 23 months (ICH); a longer latency also predicts less drug-resistant poststroke epilepsy.^37^ Late first seizures may reflect a less epileptogenic process or causes unrelated to the stroke, although shorter follow-up after late seizures may bias these estimates downward. First seizures on the day of ICH carried a higher seizure recurrence risk than those over the following week, as observed previously for day 0 acute symptomatic seizures;^8^ this may signal a particularly low seizure threshold or unmeasured factors.

Mortality broadly mirrored seizure recurrence risk inversely. This pattern is partly expected by design, because individuals who die early cannot have a recurrence, which the subdistribution approach accounts for.^18^ It may also be substantive, because lesion characteristics that increase early mortality, such as size and severity, also increase seizure risk.^38^

### Strengths and Limitations

Strengths include harmonized individual data from international cohorts covering 4 stroke subtypes, continuous competing-risk modeling of first-seizure timing, and consistent zone separation across subtypes, bootstrap resamples, and leave-one-cohort-out analyses.

This study also has limitations. First, seizure recurrence was not centrally adjudicated, ascertainment differed across cohorts, and video-electroencephalography confirmation was not systematic; early or electrographic seizures may have been underreported and nonepileptic events overreported. Differences in acute care and outpatient follow-up may bias early and late estimates, respectively. Second, follow-up was much shorter than the 10-year horizon of the ILAE definition, particularly after ICH, so 10-year risks and zone boundaries are model-based extrapolations that should be interpreted with their CIs. Zone estimates rose by a mean of 4 percentage points between 5 and 10 years after ischemic stroke and ICH, 5 points after SAH, and 16 points after CVT (eTable 11 in Supplement 1), consistent with a dynamic seizure risk.^38-40^ Third, individuals had to survive until their first seizure to enter the analysis, a survivor bias only partly addressed by competing-risk modeling. Fourth, most individuals received ASM; weighting cannot exclude residual confounding by indication, although fully untreated cohorts would be neither realistic nor ethical.^7^ Fifth, the SAH and CVT samples were smaller, with first seizures clustered early and most individuals treated; their trajectories should be regarded as hypothesis generating. Sixth, the zones rely on first-seizure timing and subtype alone and do not yet incorporate modifiers such as lesion location, size, cortical involvement, age, or status epilepticus at onset,^8-10,13,41-43^ and the LHH assumptions derive largely from populations without stroke.^16^ Seventh, no cohort was from Africa, and few were from South America or South and East Asia.

## Conclusions

In this pooled analysis, the 7-day boundary did not mark a change in seizure recurrence risk after stroke. Seizure recurrence risk rose gradually and differed by stroke subtype. Subtype-specific risk zones that combine seizure recurrence probability, competing mortality, and explicit benefit-harm assumptions could support individualized counseling and stratification of future trials but require prospective external validation with standardized seizure ascertainment before they are used to change diagnosis or treatment.

## Supporting information

Supplement 1: eMethods, eTables 1-11, eFigures 1-6, eReferences

Supplement 2: Nonauthor Collaborators

## Article Information

### Author Contribution

Drs Schubert and Galovic had full access to all of the data in the study and take responsibility for the integrity of the data and the accuracy of the data analysis.

*Concept and design:* Schubert, Galovic.

*Acquisition, analysis, or interpretation of data:* All authors.

*Drafting of the manuscript:* Schubert, Galovic.

*Critical review of the manuscript for important intellectual content:* All authors.

*Statistical analysis:* Schubert.

### Group Information

The nonauthor collaborators of the SeLECT Consortium, the International Cerebral Venous Thrombosis Consortium, the MUCH-Italy Investigators, and the ILAE Acute Symptomatic Seizures Task Force are listed in Supplement 2.

### Ethics Approval and Informed Consent

Each contributing cohort was approved by its local ethics committee or institutional review board, and written or verbal informed consent was obtained or waived according to local regulations (eTable 2 in Supplement 1). Centers contributed deidentified individual-level data. The pooled analysis was approved by the Cantonal Ethics Committee Zurich (BASEC 2024-02244).

### Trial Registration

Not applicable; this is an observational cohort study without an intervention.

### Data Availability

Deidentified participant data with a data dictionary, together with the statistical and analytic code and the study protocol, will be available after publication of the peer-reviewed article, without an end date, to qualified investigators for any purpose deemed scientifically sound and ethically approved. Access requires approval of a proposal by the lead investigator and the respective local cohort contributors, with evidence of appropriate ethical approvals. Requests should be directed to Marian Galovic, MD, PhD.

### Additional Contributions

We thank all patients and the staff of the contributing centers and consortia.

### Use of Artificial Intelligence

Claude (Anthropic; Claude Opus models accessed through Claude Code) was used from September to November 2026 for language editing, shortening the manuscript to journal length, formatting the manuscript and supplements to journal style, and preparing this preprint version. The authors reviewed and edited all output and take full responsibility for the content of this article.

