## Supplement 1: eMethods, eTables 1-11, eFigures 1-6, eReferences for "Timing of First Poststroke Seizure and Risk of Seizure Recurrence by Stroke Subtype"

This supplemental material has been provided by the authors to give readers additional information about their work.

#### eMethods

Abbreviations used in these eMethods: ASM, antiseizure medication; CI, confidence interval; CIF, cumulative incidence function; CVT, cerebral venous thrombosis; HR, hazard ratio; ICH, intracerebral hemorrhage; ILAE, International League Against Epilepsy; IPD, individual participant data; IPW, inverse-probability weighting; IS, ischemic stroke; LHH, likelihood of being helped versus harmed; NNH, number needed to harm; NNT, number needed to treat; RRR, relative risk reduction; SAH, subarachnoid hemorrhage.

##### Study design and setting

This is a retrospective, multicenter pooled analysis of individual participant data (IPD) evaluating how timing of the first seizure after stroke relates to subsequent seizure recurrence and death. The analysis combines harmonized data from SeLECT consortium centers and disease-specific collaborating cohorts contributing patients with ischemic stroke (IS), spontaneous intracerebral hemorrhage (ICH), aneurysmal subarachnoid hemorrhage (SAH), or cerebral venous thrombosis (CVT). These online methods provide additional operational detail for cohort construction, timing-window definitions, subtype-specific modeling choices, reliability diagnostics.

##### Ethics, consent, and data governance

Each contributing cohort was approved by the relevant local ethics committee or institutional review board according to local regulations. Consent requirements differed by jurisdiction and source cohort; written informed consent, verbal consent, or ethics-committee waiver of consent was used where appropriate for retrospective registry-based research. Centers contributed deidentified individual-level data required for the present analysis. Detailed center-level ethics approvals, where available, are reported in eTable 2. Per-cohort ethics and consent procedures are summarized in eTable 2.

##### Source population and eligibility

Eligible participants were adults with a first clinically confirmed seizure after ischemic stroke, spontaneous intracerebral hemorrhage, aneurysmal subarachnoid hemorrhage, or cerebral venous thrombosis. Patients were excluded if epilepsy pre-dated the stroke, if the seizure was judged unrelated to the stroke, or if first-seizure timing, follow-up time, recurrence status, or vital status was unavailable or internally inconsistent. Negative first-seizure times were excluded for IS, ICH, and SAH, but retained for CVT because seizure can precede formal CVT diagnosis and represent the presenting manifestation.

For a first seizure occurring within the first 30 days, the patient had to be followed to at least day 30 after the index event; for example, a patient whose first seizure occurred on day 16 was included only if follow-up extended to at least day 30. Early-window records without this minimum observation were excluded, so that early first seizures were not misclassified as non-recurring purely because of short follow-up. The final analysis cohort comprised 2434 patients: 931 with IS, 715 with ICH, 338 with SAH, and 450 with CVT.

| Stroke subtype | Final N | Special timing rule |
|---|---|---|
| Ischemic stroke | 931 | First-seizure day required to be >=0; early-window support rule applied. |
| Intracerebral hemorrhage | 715 | First-seizure day required to be >=0; early-window support rule applied. |
| Subarachnoid hemorrhage | 338 | First-seizure day required to be >=0; early-window support rule applied; timing-risk curve modeled with a single continuous spline (five degrees of freedom). |
| Cerebral venous thrombosis | 450 | Seizures occurring before the CVT diagnosis date (pre-index seizures) were summarized to day 0; no negative onset days enter the analysis axis. |

##### Data harmonization and verified-clean master

Center-specific source variables were harmonized into standard definitions for stroke subtype, index event, first-seizure timing, follow-up duration, seizure recurrence, death, demographics, antiseizure medication exposure, and subtype-specific clinical covariates. Final cohort membership was fixed before extended clinical variables were joined, ensuring that availability of individual covariates did not change the analysis denominator.

**Index event and seizure timing**
The index event was the date of stroke onset or, for CVT, the date of radiological CVT diagnosis. First-seizure timing was expressed as days between the index event and first documented seizure. For IS, ICH, and SAH, timing was required to be non-negative. For CVT, seizures occurring before the diagnosis date (pre-index seizures)1 were summarized to day 0, so no negative onset days enter the analysis axis.

**Outcomes**
The primary outcome was recurrent seizure after the first poststroke seizure. Death without prior recurrent seizure was treated as the competing event. The secondary competing-risk outcome was all-cause mortality after the first seizure, with seizure recurrence before death treated as the competing event. Follow-up for recurrence and mortality was counted from the first poststroke seizure.

Seizures were classified according to International League Against Epilepsy (ILAE) criteria: the index poststroke seizure and every recurrence were clinically confirmed events; acute symptomatic seizures were defined as those occurring within 7 days of the stroke, and a recurrence relevant to the epilepsy definition was an unprovoked seizure occurring thereafter. Seizure recurrence was ascertained within each contributing cohort using its standard follow-up procedures, most commonly structured in-person clinical follow-up, review of electronic medical records or hospital charts, telephone follow-up or structured interview, EEG-supported confirmation, and, for several cohorts, registry or administrative-database linkage; the procedures used by each cohort are summarized in eTable 2 (Per-cohort descriptions). To mitigate heterogeneity in ascertainment across cohorts, center-specific source variables were harmonized into standard case definitions, ambiguous cases were adjudicated, and, where cohorts performed it, source-data verification or central/adjudicated review was applied (per-cohort details in eTable 2). The influence of differing follow-up modality on the recurrence estimates was examined in leave-one-cohort-out and ascertainment-modality sensitivity analyses (eTable 8).

**Clinical covariates and subtype-specific variables**
Shared covariates in the propensity-score and outcome models were sex, age dichotomized at 65 years, and a binary early/acute first-seizure indicator (first seizure within 7 days of the index event). Age was entered continuously in the inverse-probability-weighting propensity models and dichotomized at 65 years in the outcome models. Because the early/acute first-seizure indicator is itself defined by first-seizure timing, its association with recurrence should be read as the conventional 7-day acute-symptomatic dichotomy expressed as a hazard ratio, rather than as a predictor independent of timing. Subtype-specific covariates were selected to reflect clinically plausible recurrence mechanisms and to match the adjusted models used in the figures.

| **Stroke subtype** | **Covariates used in adjusted recurrence analyses based on previous literature/ prediction models** |
|---|---|
| Ischemic stroke | Sex; age; NIHSS middle and high severity flags; large-artery atherosclerosis; cardioembolic mechanism; small-vessel disease; MCA territory involvement; cortical involvement; reperfusion treatment; early/acute first seizure.[4-8] |
| Intracerebral hemorrhage | Sex; age; NIHSS middle and high severity flags; lobar hemorrhage; hematoma volume >10 mL or continuous hematoma volume depending on the table/figure; neurosurgical intervention; early/acute first seizure.[9-11] |
| Subarachnoid hemorrhage | Sex; age; modified Rankin Scale at discharge; VASOGRADE; aneurysm treatment/surgery proxy; early/acute first seizure.[12] |
| Cerebral venous thrombosis | Sex; age; intracerebral hemorrhagic lesion at presentation; subdural hematoma; decompressive surgery; early/acute first seizure.[1] |

For baseline reporting, NIHSS categories were defined as moderate severity for NIHSS 4-10 and severe for NIHSS greater than 10.

**Handling of missing data**
Missingness was reported descriptively before model-specific imputation. For model fitting, binary covariates were imputed to the observed modal value, categorical covariates to the modal non-missing level, and numeric covariates to the observed median within the model dataset. Variables without residual variation after imputation were omitted from the affected model to avoid singular estimation. Missingness rates are reported in Table 1 and eTable 9.

**Antiseizure medication treatment and stabilized inverse probability weighting**
Antiseizure medication exposure was not randomly assigned. For adjusted cumulative incidence estimates, stabilized inverse probability treatment weights (numerator = the marginal treatment probability) were estimated with subtype-specific logistic propensity-score models, consistent with marginal structural modeling of non-randomized treatment exposure.[13,14] Binary covariates were mode-imputed and numeric covariates median-imputed before weight estimation. Weights targeted the average treatment effect, were truncated at the 1st and 99th percentiles, and were combined with Fine-Gray case weights in weighted subdistribution hazard models. Weight estimation used the WeightIt package.[15] Covariate balance before and after weighting is reported in eTable 4.

**Competing-risk cumulative incidence models**
Cumulative incidence functions were estimated with Fine-Gray subdistribution hazard models.[16,17] For recurrence analyses, recurrent seizure was the event of interest and death without prior recurrence was the competing event. For mortality analyses, death was the event of interest and recurrence before death was treated as the competing event. Between-group equality of subdistribution CIFs was assessed with Gray's test.[18]

Continuous trend curves used exact first-seizure day with natural cubic splines on log(1 + day) for non-CVT stroke subtypes.[19] For CVT, seizures occurring before the diagnosis date (pre-index seizures)[1,20] were pooled to day 0, so all stroke subtypes were modeled on the same log(1 + day) timing axis. For subarachnoid hemorrhage, unstabilized weights gave a critically low effective sample size, 12% of the nominal sample; stabilized weights (numerator = the marginal treatment probability) restored it to ≈99% of nominal, so SAH was modeled with the same single continuous spline as the other stroke subtypes, capturing both the early acute peak and the later peak. Three spline degrees of freedom were used for IS, ICH and CVT and five for SAH, whose early and late peaks required a more flexible basis; where the data could not support the basis, the degrees of freedom were reduced stepwise (CVT, with 92% of first seizures on days 0-7, was fitted with a log-linear term). The identical specification was used for the point estimates and for every bootstrap resample. All observed follow-up was used, and cumulative incidences were read at 10 years.

**Data-driven threshold search**
To evaluate whether a binary timing cutoff provided a parsimonious summary, candidate cut-points were scanned using Fine-Gray models and compared by bayesian information criterion.[21] Candidate cut-points (days 1-30 and 35-90 in 5-day steps, each group with ≥20 patients and ≥5 recurrences) were evaluated on the exact first-seizure day scale, with CVT pre-index seizures summarized to day 0, in stabilized-IPW Fine-Gray models on the final analysis cohort. The plausible range of cutoffs was defined as the span with ΔBIC ≤ 2 (strongly equivalent to the optimum) or ≤ 6 (not substantially worse), following the Kass and Raftery conventions for Bayes factor interpretation.[22] The stability of the zone boundaries was assessed with the 1000-resample bootstrap described below.[23] The threshold search was used descriptively to compare the conventional 7-day boundary with data-adapted alternatives; final timing zones were derived from the LHH framework and checked against BIC profiles, window-level estimates, and reliability diagnostics. BIC-optimal cutoffs and plausible ranges are reported in eTable 5.

**Between-subtype comparison, pooling, and resampling stability**

We tested whether the four stroke subtypes could be pooled or had to be modeled separately along three lines. First, we compared the unadjusted 10-year recurrence cumulative incidence between stroke subtypes with a four-group Gray test, and tested whether a stroke subtype effect persisted after accounting for first-seizure timing using a joint Wald test on the stroke subtype terms in a Fine-Gray model that also contained a natural cubic spline in log(1 + day) (three degrees of freedom).[17] Second, we tested whether the *shape* of the timing-recurrence trajectory differed between stroke subtypes in two complementary ways: (i) a pseudo-value generalized additive model[24] of the 10-year recurrence cumulative incidence, comparing a full model with subtype-specific smooths against a reduced model with a single common smooth (F-test); and (ii) a robust Wald test of the stroke subtype × log(1 + day) spline interaction in an inverse-probability-weighted Fine-Gray model. For these trajectory comparisons, the four stroke subtypes were compared on a common three-degree-of-freedom spline basis, with SAH as a single stratum and CVT pre-index seizures pooled at day 0, matching the modeling choices used for the curves. A significant between-subtype difference, or a significant stroke subtype × timing interaction, was taken to indicate that a single pooled curve would misrepresent at least one stroke subtype and that subtype-specific modeling was required. Third, within each stroke subtype we tested whether the 10-year recurrence cumulative incidence differed across the low-, moderate- and high-risk zones with a Gray K-sample test.

We assessed the stability of the timing-derived cutoffs with two 1000-iteration bootstrap analyses,[23] both refitting the stabilized inverse-probability-weighted Fine-Gray spline model in each resample, with window-stratified resampling for the day-0-dominated SAH and CVT strata. The first re-derived the zone boundaries in each resample with the same rule as the main analysis (entry into the moderate zone at CIF 31.6%, entry into the high-risk zone at CIF 60% and, where applicable, the last day of the high-risk zone), summarized as the bootstrap median and 95% percentile interval next to the point estimate, to quantify how sample-dependent the exact boundary day is. The second held the published zone cutoffs fixed and recomputed the within-zone recurrence cumulative incidences in each resample, recording the proportion of resamples in which the zones remained significantly distinct (Gray test) and correctly ordered (monotone), as a test of how robustly the published classification separates risk. These analyses are reported in eTable 7, eFigures 3 and 4 and Figure 4; the 95% intervals of the recurrence curves in Figures 2-4 are the percentile intervals of the first bootstrap.

**Likelihood-of-being-helped-versus-harmed framework**

To translate recurrence probabilities into clinically interpretable treatment-decision zones, we used likelihood of being helped versus harmed:[25] LHH(t) = NNH × CIF(t) × RRR. The base-case NNH was 19 and the assumed relative risk reduction was 0.25, informed by placebo-controlled antiseizure-medication trial data.[26] Under this parameterization, LHH=1.5 corresponds to a recurrence CIF of approximately 31.6%, and the ILAE 60% recurrence CIF corresponds to LHH = 2.85. The 60% level corresponds to the conventional 10-year recurrence-risk threshold used for epilepsy diagnosis after a first unprovoked seizure in an appropriate clinical context.

The framework formalizes the trade-off between the benefit and the harm of starting long-term ASM. The benefit depends on the absolute recurrence risk that can be prevented: the number needed to treat to avoid one recurrence is NNT = 1 / (CIF × RRR), where CIF is the estimated 10-year recurrence probability at a given first-seizure timing and RRR is the relative reduction in recurrence achieved by ASM. The harm of treatment is taken to be approximately independent of recurrence risk and is summarized by the number needed to harm (NNH), the number of patients treated for one to experience a clinically relevant adverse drug effect. The likelihood of being helped versus harmed is therefore LHH = NNH / NNT = NNH × CIF × RRR, which increases monotonically with the recurrence probability, so that a higher recurrence risk yields a more favorable benefit-harm balance for treatment. Inverting the identity gives the recurrence level corresponding to any LHH value, CIF = LHH / (NNH × RRR); we used this to convert the two decision thresholds into recurrence-CIF cutoffs (eTable 3).

Base-case parameters were taken from placebo-controlled antiseizure-medication data: a relative risk reduction RRR = 0.25 in seizure recurrence and a number needed to harm NNH = 19 for clinically relevant adverse effects. The two LHH boundaries were chosen to be clinically rather than statistically motivated. The lower boundary, LHH = 1.5, is the recurrence level at which the chance of benefit first clearly outweighs the chance of harm (a 3:2 ratio); under the base case this is CIF = 1.5 / (19 × 0.25) = 31.6%. The upper boundary was fixed at the ILAE diagnostic threshold for epilepsy after a single unprovoked seizure, a ≥60% 10-year recurrence probability; at CIF = 60% the LHH is 19 × 0.60 × 0.25 = 2.85. First-seizure days were accordingly classified as low (CIF < 31.6%; LHH < 1.5), moderate (31.6% ≤ CIF < 60%), or high recurrence risk (CIF ≥ 60%; LHH ≥ 2.85). Because the boundaries follow directly from RRR and NNH, they can be recomputed for any alternative treatment-effect or harm assumption; eTable 3 reports the recurrence-CIF level of the lower boundary and the LHH implied by the fixed 60% upper boundary across a grid of alternative RRR and NNH values.

**Derivation of the base-case parameters (RRR and NNH).** The relative risk reduction (RRR = 0.25) represents an approximately 25% relative reduction in the risk of a further seizure with antiseizure-medication (ASM) treatment. This is the order of effect observed in randomized controlled trials of immediate versus deferred ASM after a first seizure or early epilepsy: in the Multicentre Study of Early Epilepsy and Single Seizures (MESS), immediate treatment prolonged the time to a further seizure with a hazard ratio of about 1.4, corresponding to roughly a 25-30% relative reduction.[27] The number needed to harm (NNH = 19) is the number of patients treated for one additional treatment discontinuation due to adverse effects. It is drawn from the only harmonized synthesis of placebo-controlled ASM benefit-harm data,[26] as recently tabulated for LHH calculation by Brigo.[28] In that synthesis the NNH for withdrawal due to adverse effects was 19 for several widely used ASMs (for example pregabalin and lacosamide) and 26 (95% CI 21-38) pooled across all agents; we adopted the lower, more conservative value of 19 so as not to overstate the net benefit of treatment. Both parameters are made explicit and can be varied: eTable 3 reports the resulting recurrence-CIF boundaries across a grid of alternative RRR and NNH values and shows that the low/moderate/high structure is preserved across plausible inputs.

**Rationale for the LHH boundaries.** We first anchored the high-risk boundary to the established threshold at which the International League Against Epilepsy (ILAE) considers a single seizure sufficient to diagnose epilepsy, a ≥60% 10-year risk of a further seizure.[3] Under the base case this corresponds to LHH = 19 × 0.60 × 0.25 = 2.85. We then set the low-risk boundary at half this value, LHH = 1.5, the point at which a patient is 1.5 times (a 3:2 ratio) more likely to be helped than harmed, corresponding to CIF = 1.5 / (19 × 0.25) =

31.6%. Placing the lower boundary at half the epilepsy-defining threshold deliberately builds in a safety margin: below LHH 1.5 the expected benefit of long-term ASM is small and inconsistent, whereas the moderate zone (LHH 1.5-2.85) captures the range in which benefit is plausible but uncertain and treatment should be individualized through shared decision-making. The precise value is a pragmatic, rounded choice rather than a hard cutoff; neighboring values such as LHH 1.3 or 1.7 (≈27% and ≈36% 10-year recurrence) would shift the low/moderate boundary by only a few percentage points and leave the three-zone structure unchanged (eTable 3).

Recurrence-risk zones were derived directly from each stroke subtype's smoothed recurrence curve using the LHH framework: a given first-seizure day was classified as low risk where LHH<1.5 (10-year recurrence CIF <31.6%), moderate where LHH≥1.5 but CIF<60%, and high where CIF≥60% (LHH ≥2.85, the ILAE epilepsy threshold). Because a curve can cross 60% and later fall back below it, these instantaneous categories need not be contiguous in time, most evident in subarachnoid hemorrhage, whose curve peaks early, declines, and rises again; the IS and ICH curves also fall back below 60% after about 16 and 23 months. Zone derivation was capped at day 1,000, beyond which too few first seizures were observed to define further transitions; the zone active at day 1000 was carried forward. For each stroke subtype, the calendar days at which the spline-smoothed recurrence curve crossed the 31.6% and 60% CIF thresholds defined the boundaries of the risk zones.

| Stroke subtype | Low risk zone | Moderate risk zone | High risk zone |
|---|---|---|---|
| Ischemic stroke | Days 0-14 | Days 15-70 and ≥492 | Days 71-491 |
| Intracerebral hemorrhage | Days 1-8 | Day 0, days 9-40 and ≥692 | Days 41-691 |
| Subarachnoid hemorrhage | Day 0 | Day 1 and ≥4 | Days 2-3 |
| Cerebral venous thrombosis | - | Days 0-8 | Day 9 onward |

**Aalen-Johansen reliability analyses**
Aalen-Johansen cumulative incidence curves[29] were used as an assumption-light reliability check for model-based Fine-Gray recurrence estimates. Curves were estimated separately by stroke subtype and instantaneous risk category. Reliability was summarized at 1, 2, and 5 years using nominal sample size, effective sample size (ESS),[30] number at risk, and confidence-interval half-width.

The reliability figure used four pre-specified diagnostic tiers, reliable, borderline, unstable, and critical, intended to flag zone-level estimates with limited statistical support rather than to serve as formal inferential cutoffs. Tiers were defined on three criteria: effective sample size as a fraction of zone N (greater than 50%, 25-50%, 15-25%, and less than 15%), the number at risk (at least 20, 10-19, 5-9, and fewer than 5), and the half-width of the 10-year recurrence confidence interval (less than 10, 10-15, 15-20, and at least 20 percentage points). These thresholds are pragmatic,[30,31] were chosen a priori to make the displayed precision interpretable, and do not affect the point estimates or the formal statistical tests.

**Software and reproducibility**
Analyses were performed in R version 4.5.2. Numerical values, curves, confidence intervals, zone assignments, and tables were generated analytically before final graphical formatting; visual formatting did not alter underlying estimates.

#### eTables

**eTable 1.** *Per-center patient contributions*

| | | | **Enrolled (N)** | **Enrolled (N)** | **Enrolled (N)** | **Enrolled (N)** | **Enrolled (N)** | **Final analysis (N)** | **Final analysis (N)** | **Final analysis (N)** | **Final analysis (N)** | **Final analysis (N)** |
|---|---|---|---|---|---|---|---|---|---|---|---|---|
| Center/ cohort | Country | Stroke subtypes | IS | ICH | SAH | CVT | Total | IS | ICH | SAH | CVT | Total |
| Zurich | Switzerland | IS, ICH, SAH, CVT | 163 | 149 | 117 | 68 | 497 | 140 | 129 | 111 | 55 | 435 |
| MUCH Italy | Italy | ICH | - | 316 | - | - | 316 | - | 284 | - | - | 284 |
| NCVC | Japan | IS, ICH | 114 | 104 | - | - | 218 | 72 | 74 | - | - | 146 |
| AMC CVT patients | Netherlands | CVT | - | - | - | 119 | 119 | - | - | - | 111 | 111 |
| Cleveland | USA | IS | 57 | 44 | 18 | - | 119 | 56 | 42 | 15 | - | 113 |
| Vall d'Hebron | Spain | SAH | - | - | 116 | - | 116 | - | - | 100 | - | 100 |
| Athens | Greece | IS, ICH, SAH | 78 | 17 | 8 | - | 103 | 75 | 16 | 7 | - | 98 |
| Homburg | Germany | IS, SAH | 84 | - | 17 | - | 101 | 78 | - | 15 | - | 93 |
| KSSG | Switzerland | IS | 97 | - | - | - | 97 | 92 | - | - | - | 92 |
| China | China | ICH | - | 100 | - | - | 100 | - | 91 | - | - | 91 |
| Modena | Italy | IS | 87 | - | - | - | 87 | 87 | - | - | - | 87 |
| Udine | Italy | IS, ICH, SAH | 23 | 61 | 16 | - | 100 | 22 | 55 | 8 | - | 85 |
| Bern | Switzerland | CVT | - | - | - | 88 | 88 | - | - | - | 74 | 74 |
| Gothenburg CVT patients | Sweden | CVT | - | - | - | 68 | 68 | - | - | - | 63 | 63 |
| Fleni - Buenos Aires | Argentina | IS | 60 | - | - | - | 60 | 59 | - | - | - | 59 |
| Malaysia | Malaysia | IS, ICH, SAH | 51 | 7 | 1 | - | 59 | 47 | 5 | 0 | - | 52 |
| Bellvitge | Spain | SAH | - | - | 57 | - | 57 | - | - | 50 | - | 50 |
| Barcelona | Spain | IS, CVT | 46 | - | - | 6 | 52 | 40 | - | - | 6 | 46 |
| Linz | Austria | IS | 74 | - | - | - | 74 | 41 | - | - | - | 41 |
| Thessaloniki | Greece | IS, ICH | 37 | 4 | - | - | 41 | 37 | 4 | - | - | 41 |
| Innsbruck | Austria | IS | 39 | - | - | - | 39 | 37 | - | - | - | 37 |
| Lisbon | Portugal | IS | 38 | - | - | - | 38 | 31 | - | - | - | 31 |
| Istanbul CVT patients | Turkey | CVT | - | - | - | 29 | 29 | - | - | - | 27 | 27 |
| Helsinki CVT patients | Finland | CVT | - | - | - | 48 | 48 | - | - | - | 26 | 26 |
| Berlin | Germany | ICH, SAH | - | 16 | 11 | - | 27 | - | 15 | 10 | - | 25 |
| Erasme Hospital - Brussels | Belgium | SAH | - | - | 29 | - | 29 | - | - | 22 | - | 22 |
| Mexico City CVT patients | Mexico | CVT | - | - | - | 23 | 23 | - | - | - | 21 | 21 |
| Hamadan CVT patients | Iran | CVT | - | - | - | 20 | 20 | - | - | - | 17 | 17 |
| Lisbon HSM CVT patients | Portugal | CVT | - | - | - | 14 | 14 | - | - | - | 14 | 14 |
| Lisbon HGO CVT patients | Portugal | CVT | - | - | - | 13 | 13 | - | - | - | 11 | 11 |
| Muenster | Germany | IS | 32 | - | - | - | 32 | 9 | - | - | - | 9 |
| Palermo CVT patients | Italy | CVT | - | - | - | 9 | 9 | - | - | - | 9 | 9 |
| Colombia | Colombia | IS | 26 | - | - | - | 26 | 8 | - | - | - | 8 |
| Costa Rica CVT patients | Costa Rica | CVT | - | - | - | 11 | 11 | - | - | - | 8 | 8 |

| | | | | | | | | | | | | |
|---|---|---|---|---|---|---|---|---|---|---|---|---|
| Homburg CVT patients | Germany | CVT | - | - | - | 6 | 6 | - | - | - | 5 | 5 |
| Australia JHH CVT patients | Australia | CVT | - | - | - | 4 | 4 | - | - | - | 3 | 3 |
| South Limburg Cohort | Netherlands | ICH | - | 138 | - | - | 138 | - | 0 | - | - | 0 |
| Total | | | 1106 | 956 | 390 | 526 | 2978 | 931 | 715 | 338 | 450 | 2434 |

Per-center enrolled (raw) and final-analysis (eligible) counts by stroke subtype. Final-analysis columns reflect the final eligibility cascade (n=2434: IS 931, ICH 715, SAH 338, CVT 450). N (enrolled/final) sums to 2978/2434; the South Limburg cohort (row 37) contributed no eligible individuals.

Abbreviations. CVT = cerebral venous thrombosis; ICH = intracerebral hemorrhage; IS = ischemic stroke; SAH = subarachnoid hemorrhage.

**eTable 2.** ***Per-cohort descriptions***

*Design, eligibility, seizure ascertainment, follow-up, ethics/consent, and source for each contributing center (eTable 1). Cerebral venous thrombosis (CVT) registries contributed collectively through the International CVT Consortium (ICVTC); their shared methods are given in the note beneath the table.*

| # | Center/cohort (PI; country) | Stroke subtypes; N (enrolled/final); design & period | Eligibility; index-seizure definition; ascertainment & follow-up | Ethics / consent; source |
|---|---|---|---|---|
| 1 | Zurich<br>PI: Kai Michael Schubert, Marian Galovic<br>Switzerland | IS, ICH, SAH, CVT<br>N 497/435<br>Coordinating SeLECT cohort, tertiary referral center (Zurich). | Incl: Adults ≥18 years with a documented ischemic stroke (IS), intracerebral hemorrhage (ICH), subarachnoid hemorrhage (SAH), or cerebral venous thrombosis (CVT), and a first documented acute symptomatic or remote symptomatic seizure after the qualifying stroke; availability of relevant clinical data and longitudinal follow-up. Excl: Age <18 years; previous epilepsy or seizures predating the qualifying stroke; seizures attributable to another acute or chronic epileptogenic condition rather than the index stroke; missing or incomplete medical records; insufficient follow-up or loss to follow-up shortly after the index event. Index: First documented poststroke seizure; seizures within 7 days of stroke onset were classified as acute symptomatic and those >7 days as remote symptomatic, with finer timing intervals according to the harmonized SeLECT-RS definitions. Ascertainment: Retrospective review of routine clinical records, including neurological documentation, imaging, and EEG findings when available. Follow-up: From the index seizure until seizure recurrence, death, or last available clinical follow-up. | Ethics: Cantonal Ethics Committee Zurich (BASEC 2024-02244); General Consent or approved Art. 34 HRA exemption where applicable. Source: SeLECT-RS protocol v3.0 and ethics approval. |
| 2 | MUCH-Italy (Multicenter Study on Cerebral Hemorrhage in Italy)<br>Coordinator: Alessandro Pezzini<br>Italy | ICH<br>N 316/284<br>prospective, multicenter consortium of 12 Italian neurological centers; January 1, 2002 - July 31, 2014. | Incl: first-ever acute spontaneous ICH - no history of previous stroke Excl: secondary causes of ICH - previous history of epilepsy<br>Index: clinical-EEG definition Ascertainment: Structured in-person clinical follow-up, Review of electronic medical records / hospital charts, Telephone follow-up / structured interview Follow-up: In person follow-up evaluations were conducted at 3 months and then annually | Ethics: MUCH-Italy received approval from the EC of Brescia University Hospital Source: authorship & cohort questionnaire. |
| 3 | Japan (NCVC, Osaka)<br>PI: Kazuki Fukuma, Masafumi Ihara<br>Japan | IS, ICH<br>N 218/146<br>Single-center retrospective observational cohort; January 1, 2012 - December 31, 2023. | Incl: Adults (≥18 years) with acute, first-ever ischemic stroke or spontaneous intracerebral hemorrhage confirmed by CT or MRI, admitted within 3 days of stroke onset, and diagnosed with poststroke seizures. Excl: History of epilepsy or unprovoked seizures before stroke, severe traumatic brain injury, intracranial tumors, cerebral venous thrombosis, or previous brain surgery. Index: The first seizure after the qualifying stroke, according to the predefined SeLECT-RS study definitions. Ascertainment: Structured in-person clinical follow-up, Review of electronic medical records / hospital charts, EEG-supported confirmation Follow-up: Seizure recurrence was identified through outpatient follow-up and review of electronic medical records. Follow-up continued until the last available clinical visit. The median follow-up duration was 23.1 months (IQR 2.9 to 62.7) for IS and 7.6 months (IQR 1.2 to 42.7) for ICH. | Ethics: Approved by the Institutional Review Board of the National Cerebral and Cardiovascular Center (IRB No. M30-051-5). Written informed consent was waived because of the retrospective observational study design. Source: authorship & cohort questionnaire. |
| 4 | Amsterdam University Medical Centers (AMC)<br>PI: Jonathan M Coutinho<br>Netherlands | CVT<br>N 119/111<br>CVT registry (ICVTC) | See ICVTC note below table. | Ethical Review Committee of the Amsterdam UMC decided study is not subject to the Dutch Medical Research Involving Human Subjects Act |
| 5 | Cleveland<br>PI: Vineet Punia<br>USA | IS, ICH, SAH<br>N 119/113<br>Retrospective, prospectively maintained stroke & cEEG database; 1 Apr 2012-31 Mar 2018. | Adults with acute stroke undergoing cEEG within 7 days of last-known-well; excluded prior epilepsy. PSE = clinical seizures after discharge; cEEG + EMR review (blinded); EEG per ACNS/Salzburg. | IRB approval (retrospective). Source: SeLECT-EEG (Ann Neurol 2025), eMethods 'USA'. |
| 6 | Vall d'Hebron / Barcelona<br>PI: Laura Abraira<br>Spain | SAH<br>N 116/100<br>Multicenter prospective longitudinal cohort, six | Adults ≥18 with acute IS or hemorrhagic stroke; neuroimaging-confirmed. Excluded no-consent, >6 h onset-to-ER, prior epilepsy, TIA, AVM, subdural hematoma. Clinical monitoring; chart review + | Written informed consent (multicenter protocol). Source: $SeLECT_{2.0}$ (JAMA Neurol 2023), eMethods 'Spain'. |

| | | | | |
|---|---|---|---|---|
| | | Catalan hospitals; Aug 2012-Nov 2013. | telephone interviews. | |
| 7 | Athens<br>PI: Panagiota-Eleni Tsalouchidou<br>Greece | IS, ICH, SAH<br>N 103/98<br>retrospective registry; 2011-2025 | Incl: Adult patients with ischemic stroke, intracerebral hemorrhage (ICH), or subarachnoid hemorrhage (SAH) who experienced at least one acute symptomatic or remote symptomatic seizure after stroke, with available clinical and follow-up data on seizure recurrence and antiseizure medication treatment. Excl: Patients without documented poststroke seizures, those with insufficient clinical or follow-up data, or those with seizures attributable to causes other than stroke (e.g., pre-existing epilepsy or other acute neurological/metabolic conditions). Index: The index poststroke seizure was defined as the first documented clinical seizure occurring after ischemic stroke, intracerebral hemorrhage (ICH), or subarachnoid hemorrhage (SAH), irrespective of whether it was classified as an acute symptomatic or remote symptomatic seizure. Seizures were identified through review of electronic medical records and, when available, structured in-person clinical follow-up or telephone interviews. The diagnosis was confirmed by the treating neurologist, with EEG findings used to support the diagnosis when available. The date of the first poststroke seizure was recorded as the index event. Ascertainment: Structured in-person clinical follow-up, Review of electronic medical records / hospital charts, Telephone follow-up / structured interview, EEG-supported confirmation Follow-up: Seizure recurrence was defined as any subsequent unprovoked seizure occurring after the first documented poststroke seizure. Recurrence was ascertained through review of electronic medical records, outpatient clinic documentation, hospital records, and, when available, structured in-person follow-up or telephone interviews. Information on seizure recurrence, the timing of recurrent seizures, antiseizure medication treatment, and follow-up duration was recorded whenever available. Follow-up time was defined as the interval from the first poststroke seizure to the last available clinical follow-up. | Ethics: Institutional Review Board (IRB): Scientific Council of the University General Hospital "Attikon", Athens, Greece, following a favorable opinion from the Hospital Bioethics Committee. Retrospective observational cohort based on routinely collected clinical data. Source: authorship & cohort questionnaire. |
| 8 | Homburg<br>PI: Piergiorgio Lochner<br>Germany | IS, SAH<br>N 101/93<br>retrospective registry, multicenter consortium; Jan 2012- Dez 2022- Follow-up 2025 | Incl: According to select-RS Ischemic stroke: Ischemic stroke cases First-time acute or remote symptomatic seizure, with as precise documentation of the seizure time (date) as possible Information on whether another seizure occurred afterward (yes/no), the timing of any recurrent seizures, preferab…<br>Index: according to Select-RS (Excel File) Ascertainment: Structured in-person clinical follow-up, Registry or administrative-database linkage, EEG-supported confirmation Follow-up: Review of electronic medical records / hospital charts | Ethics: 142/18 and 40/20 Universität Saarlandes |
| 9 | St. Gallen (KSSG)<br>PI: Dominik Zieglgänsberger<br>Switzerland | IS<br>N 97/92<br>Retrospective cohort, tertiary referral center (KSSG); 1 Jan 2002-31 Dec 2008. | Consecutive adults ≥18 with acute first-ever neuroimaging-confirmed IS. Excluded TIA, prior stroke, primary hemorrhage, prior seizures, re-infarction, epileptogenic comorbidities. Structured telephone interview (median 28 mo); positive answers → face-to-face + EEG. | Per original SeLECT approval. Source: SeLECT 2018 (Lancet Neurol) / SeLECT$_{2.0}$ (JAMA Neurol 2023), eMethods ‘Switzerland (1)’. |
| 10 | Qingdao (China)<br>PI: Yan Wang<br>China | ICH<br>N 100/91<br>Retrospective cohort (Affiliated Hospital of Qingdao University; Qingdao Municipal Hospital); 2014-2017 / 2015-2017. | Consecutive adults ≥18 with acute neuroimaging-confirmed ICH within 14 days; CT on admission. Excluded tumor, TBI, SAH, hemorrhagic transformation, prior seizures/AED. Early ≤7 d / late >7 d. Telephone follow-up (validated questionnaire) + epilepsy-clinic EEG; ILAE diagnosis. | Institutional approval. Source: document provided by Y. Wang (Qingdao/LANE cohort). |
| 11 | Modena<br>PI: Stefano Meletti<br>Italy | IS<br>N 87/87<br>retrospective registry; 2014-2022 | Incl: Consecutive adults with acute first-ever neuroimaging-confirmed ischemic stroke who were admitted at the Stroke Unit of the Ospedale Civile Baggiovara (Modena, Italy) Excl: Subjects with transient ischemic attack (TIA), previous history of stroke, previous history of seizures, recurrent stroke during follow-up were excluded. Transient ischemic attack was defined as as a transient episode of neurological dysfunction caused by focal brain ischemia, without evidence of acute | Ethics: The scientific advisory board of our Institution approved the research protocol according to local regulations and the retrospective analysis of patients’ data. The SITS-ISTR dataset was approved by the local ethics committee. Informed consent was obtained from all individual participants included in the registry. Source: authorship & cohort |

| | | | | |
|---|---|---|---|---|
| | | | infarction. We also excluded patients with comorbidities known to have a significant risk to develop seizures (“epileptogenic comorbidities”), e.g., alcohol or drug abuse, previous brain lesions as intracranial tumors, cerebral venous thrombosis, cerebral bleeding, severe traumatic brain injury, supratentorial brain surgery, and cerebral arteriovenous malformations. Finally, subjects who were lost to follow-up were excluded; data were not available for subjects who died within 30 days from the index stroke. Index: Seizures that occurred within 7 days of stroke onset were considered as acute symptomatic seizures and those that occurred after 7 days were considered as unprovoked seizures. Poststroke epilepsy was diagnosed as the occurrence of one or more unprovoked seizures (i.e., seizures occurring beyond 7 days of stroke onset) during the follow-up6 according to the current operational definition of epilepsy. Ascertainment: Structured in-person clinical follow-up, Review of electronic medical records / hospital charts Follow-up: By internal protocol, patients undergo outpatient follow-up at 3 at 12 months; subsequent phone interviews are performed at 24 and 36 months. Of note, all patients with acute neurological symptoms (including seizures) are registered in the same hospital information system. Therefore, follow-up data were acquired by the computerized hospital chart review, outpatients’ visits, and telephone interviews, and were updated to 31st of December 2024. The diagnosis of PSE was assessed and confirmed by epileptologists of the Modena comprehensive epilepsy center, who could acquire information about the medical history of patients. | questionnaire. |
| 12 | Udine<br>PI: Gian Luigi Gigli, Giovanni Merlino<br>Italy | IS, ICH, SAH<br>N 100/85<br>Retrospective cohort, Stroke Unit, Udine University Hospital; Jul 2008-Sep 2024. | Adults ≥18 with neuroimaging-confirmed stroke; EEG during hospitalization. Excluded TIA, prior epilepsy, severe TBI, brain surgery, tumors, other epileptogenic conditions. TOAST classification; outpatient follow-up; mean ≈73 mo. | Institutional approval (retrospective). Source: SeLECT-EEG (Ann Neurol 2025), eMethods ‘Italy’. |
| 13 | Inselspital Bern University Hospital, Bern<br>PI: Mirjam R Heldner<br>Switzerland | CVT<br>N 88/74<br>CVT registry (ICVTC) | See ICVTC note below table. | The study was approved by the local ethics committee of the canton of Bern/Switzerland. |
| 14 | Sahlgrenska University Hospital, Gothenburg<br>PI: Katarina Jood<br>Sweden | CVT<br>N 68/63<br>CVT registry (ICVTC) | See ICVTC note below table. | Permission granted via the Swedish Ethical Review Authority in accordance with the Swedish Ethical Review Act (SFS 2003:460). |
| 15 | Fleni - Buenos Aires<br>PI: Matías Alet<br>Argentina | IS<br>N 60/59<br>Retrospective registry, multicenter consortium; Jan 2012 - Dec 2020 | Incl: Adults (≥18 years) with first-ever acute ischemic stroke who experienced at least one clinically confirmed seizure defined per ILAE definitions. Excl: Pre-existing seizures/epilepsy, seizures of nonstroke etiology, or follow-up <30 days. Index: ILAE classification of poststroke seizures, early and late seizures using a 7-d threshold; seizures occurring within and after 7 d of stroke onset were classified as early and late seizures, respectively. Ascertainment: Structured in-person clinical follow-up, Review of electronic medical records / hospital charts, Registry or administrative-database linkage, EEG-supported confirmation Follow-up: Seizure recurrence was ascertained according to the internal protocol with: In-person clinical follow-up Review of electronic medical records Registry or administrative-database linkage | Ethics: Participation in the study was approved by the Ethics Committee of Fleni. An exemption from informed consent was granted as it was an observational and retrospective study. Source: authorship & cohort questionnaire. |
| 16 | Malaysia (Kuala Lumpur)<br>PI: Ching Soong Khoo<br>Malaysia | IS, ICH, SAH<br>N 59/52<br>Retrospective registry; December 2011 - April 2023 | Incl: Patients with seizure onset after ischemic stroke; Patients with seizure onset after intracranial hemorrhage; Patients with seizure onset after subarachnoid hemorrhage. Excl: Patients with seizure onset before stroke; Patients with prior diagnosis of epilepsy; Patients with seizures seconda…<br>Index: By history, assessment, imaging, and EEG | Ethics: Waiver Source: authorship & cohort questionnaire. |

| | | | | |
|---|---|---|---|---|
| | | | Ascertainment: Structured in-person clinical follow-up, Review of electronic medical records / hospital charts, Telephone follow-up / structured interview Follow-up: Review of electronic medical records / hospital charts | |
| 17 | Bellvitge<br>Spain | SAH<br>N 57/50<br>Multicenter Catalonia cohort (Bellvitge site), SAH; Aug 2012-Nov 2013. | As for the Catalonia cohort; SAH cases. Clinical monitoring; chart review + telephone interviews. | Written informed consent. Source: SeLECT 2.0 (JAMA Neurol 2023), eMethods 'Spain'. |
| 18 | Barcelona<br>PI: Laura Abraira; CVT cases from Hospital de la Santa Creu i Sant Pau (PI: Daniel Guisado-Alonso)<br>Spain | IS, CVT<br>N 52/46<br>Multicenter prospective longitudinal cohort, six Catalan hospitals; Aug 2012-Nov 2013. CVT cases contributed via the ICVTC; see note below table. | Adults ≥18 with acute IS or hemorrhagic stroke; neuroimaging-confirmed. Excluded no-consent, >6 h onset-to-ER, prior epilepsy, TIA, AVM, subdural hematoma. Clinical monitoring; chart review + telephone interviews. | Written informed consent (multicenter protocol). Source: SeLECT 2.0 (JAMA Neurol 2023), eMethods 'Spain'. |
| 19 | Linz<br>PI: Raimund Helbok, Joachim Gruber<br>Austria | IS<br>N 74/41<br>retrospective; Jan 2011 - Dec 2017 | Incl: All patients who had been hospitalized due to an acute ischemic stroke from 2011 to 2017 who had undergone mechanical thrombectomy for intracranial large vessel occlusion were included into the study, provided they had a post-interventional brain MRI Excl: without a postinterventional brain MRI, younger than 18 years at stroke onset, who had previ- ous epileptic seizures or pre-existing brain lesions and those who died within 24 h of stroke onset Index: Poststroke seizures are divided into early versus late seizures (≤ 7 vs. > 7 days after the ictus) Ascertainment: Structured in-person clinical follow-up, Review of electronic medical records / hospital charts, Telephone follow-up / structured interview, EEG-supported confirmation Follow-up: Review of electronic medical records / hospital charts, Telephone follow-up / structured interview | Ethics: Ethics Committee of the Medical Faculty of Johannes Kepler University (JKU), 1319/2025 Source: authorship & cohort questionnaire. |
| 20 | Thessaloniki<br>PI: Vasilios K Kimiskidis<br>Greece | IS, ICH<br>N 41/41<br>Retrospective registry; Jan 2021-Dec 2025 | Incl: 1)Adults with a first clinically identified and confirmed seizure after ischemic stroke, spontaneous intracerebral hemorrhage, aneurysmal subarachnoid hemorrhage, or cerebral venous thrombosis. 2) Minimum observation period of 30 days after the index event Excl: 1) Epilepsy pre-dated the stroke 2) Seizure unrelated to the stroke per the investitators judgment 3) Unavailable or inconsistent first-seizure timing, follow-up time, recurrence status, or vital status Index: The first poststroke seizure was identified and confirmed clinically with EEG support obtained within 7 days from brain insult Ascertainment: Structured in-person clinical follow-up, Review of electronic medical records / hospital charts, Telephone follow-up / structured interview, EEG-supported confirmation Follow-up: In-person and telephone follow-up | Ethics: Written Source: authorship & cohort questionnaire. |
| 21 | Innsbruck<br>PI: Luisa Delazer<br>Austria | IS<br>N 39/37<br>prospective single-center cohort; Dec 2020 - Jan 2025 | Incl: Ischemic stroke, age >18 years, stroke symptom onset <30 days before admission Excl: (1) prior history of epileptic seizures, (2) primary hemorrhagic stroke, (3) potentially epileptogenic comorbidities (e.g., alcohol/drug abuse, intracranial tumor, cerebral venous thrombosis, severe traumatic brain injury, brain surgery, meningitis, cerebral arteriovenous malformations, large aneurysms, vasculitis, or hydrocephalus). Index: clinical assessment by a neurologist and confirmed by review of the medical records, including witness descriptions, neurological evaluation, EEG findings when available, and neuroimaging to exclude alternative causes Ascertainment: Structured in-person clinical follow-up, Review of electronic medical records / hospital charts Follow-up: standardized follow-up visits (3 and 12 months, then annually), supplemented by review of hospital records and telephone interviews. All suspected events were adjudicated by neurologists based on clinical documentation, with EEG and neuroimaging reviewed when available. | Ethics: patients gave written informed consent. clinicaltrials.gov number NCT04582825, ethics vote Innsbruck EK-Nr: 1182/2020) Source: authorship & cohort questionnaire. |

| | | | | |
|---|---|---|---|---|
| 22 | Lisbon<br>Portugal | IS<br>N 38/31<br>Prospective longitudinal cohort, university-hospital stroke unit; 24-mo enrollment, 12-mo follow-up. | Adults with neuroimaging-confirmed anterior-circulation IS; mRS ≤1; NIHSS ≥4; no prior epilepsy/head trauma/brain surgery. Blinded phone interview at 6 mo; clinical appointment at 12 mo; EEG within 72 h. | Institutional approval; consent per protocol. Source: SeLECT 2.0 / SeLECT-EEG 'Portugal' (PI: C. Bentes). |
| 23 | Istanbul University Hospital, Istanbul<br>PI: Nilüfer Yesilot<br>Turkey | CVT<br>N 29/27<br>CVT registry (ICVTC) | See ICVTC note below table. | A local ethics committee application was not made for retrospective collection of observational data |
| 24 | Helsinki University Hospital, Helsinki<br>PI: Jukka Putaala<br>Finland | CVT<br>N 48/26<br>CVT registry (ICVTC) | See ICVTC note below table. | The study was granted research permission by a local ethical committee (22.3.2018 decision reference number HUS/125/2018). |
| 25 | Berlin<br>PI: Jakob I. Doerrfuss<br>Germany | ICH, SAH<br>N 27/25<br>retrospective registry; Jan 2010 - Sep 2025 | Incl: Adult patients ≥18 with acute ICH or SAH and ≥ 1 acute symptomatic or unprovoked seizure Excl: Insufficient data, prior epilepsy. Index: acute symptomatic seizure: seizure within 1 week of stroke, unprovoked seizure: seizure >1 week after stroke Ascertainment: Structured in-person clinical follow-up, Review of electronic medical records / hospital charts Follow-up: ascertained through clinical assessment during outpatient visits, follow-up intervals were not fixed and varied between patients. | Ethics: Approval of local ethics committee (EA4/088/25) Source: authorship & cohort questionnaire. |
| 26 | Erasme Hospital - Brussels<br>PI: Nicolas Gaspard<br>Belgium | SAH<br>N 29/22<br>retrospective registry; 2016-2022 | Incl: Continuous EEG per clinical indications (ACNS guidelines, IS with NIHSS>8, Excl: Life expectancy <48h<br>Index: clinicians' judgment only OR video-EEG confirmation (if occurring during video-EEG)<br>Ascertainment: Review of electronic medical records / hospital charts, Registry or administrative-database linkage Follow-up: vascular neurology outpatient follow-up and epilepsy follow-up, if indicated | Ethics: Comité d'Ethique Hospitalo-Facultaire Erasme. Waiver of consent. Source: authorship & cohort questionnaire. |
| 27 | National Institute of Neurology and Neurosurgery Manuel Velasco Suarez, Mexico-City<br>PI: Antonio Arauz<br>Mexico | CVT<br>N 23/21<br>CVT registry (ICVTC) | See ICVTC note below table. | Permission granted by the Ethics committee of the National Institute of Neurology in Accordance with Mexican health regulations. |
| 28 | Sina Hospital, Hamadan<br>PI: Masoud Ghiasian<br>Iran | CVT<br>N 20/17<br>CVT registry (ICVTC) | See ICVTC note below table. | The ethics committee of the university decided to collect information after receiving patient consent. |
| 29 | Hospital de Santa Maria/CHULN, Lisbon<br>PI: Diana Aguiar de Sousa<br>Portugal | CVT<br>N 14/14<br>CVT registry (ICVTC) | See ICVTC note below table. | Permission granted by the Ethical committee of Centro Hospitalar Lisboa Norte (CHLN) and Centro Académico de Medicina de Lisboa (CAML), Lisbon, Portugal |
| 30 | Hospital Garcia de Orta, Almada<br>PI: Miguel Rodrigues<br>Portugal | CVT<br>N 13/11<br>CVT registry (ICVTC) | See ICVTC note below table. | Permission granted by the Ethics committee of Hospital Garcia de Orta in accordance with Portuguese health regulations |
| 31 | Muenster<br>Germany | IS<br>N 32/9<br>Retrospective cohort, tertiary referral center; 1 Jan 2003-31 Mar 2010. | Adults with first-ever hemispheric stroke. Excluded recurrent stroke, infratentorial-only, hemorrhage, TIA, CVT, in-hospital death, lost to follow-up. Structured telephone interview; median follow-up 23 mo. | Institutional approval. Source: SeLECT 2.0 (JAMA Neurol 2023), eMethods 'Germany (2)'. |
| 32 | University Hospital Policlinico Paolo Giaccone, Palermo<br>PI: Valentina Arnao<br>Italy | CVT<br>N 9/9<br>CVT registry (ICVTC) | See ICVTC note below table. | The Ethical local Committee (Palermo) takes account of this study. According to Italian legislation, non-interventional studies do not require ethical approval. |
| 33 | Colombia (Bogota) | IS | Adults ≥18 with acute IS, neurologist-diagnosed | Per SeLECT 2.0 approval. Source: |

| | | | | |
|---|---|---|---|---|
| | PI: Luis Carlos Mayor-Romero, María Paula Zafra-Sierra, Carolina Ferreira-Atuesta<br>Colombia | N 26/8<br>Retrospective cohort, university-hospital stroke unit; 1 Jan 2014-31 Dec 2019. | and neuroimaging-confirmed. Excluded primary hemorrhage, TIA, prior seizures, epileptogenic comorbidities. Medical-record review; mean follow-up ≈21 mo. | SeLECT 2.0 (JAMA Neurol 2023), eMethods 'Colombia'. |
| 34 | Hospital Dr. R.A. Calderón Guardia, CCSS, San José<br>PI: Miguel Barboza<br>Costa Rica | CVT<br>N 11/8<br>CVT registry (ICVTC) | See ICVTC note below table. | Institutional IRB from the Caja Costarricense del Seguro Social, CLOBI-HRACG, in accordance with national research policies and Helsinki Declaration. |
| 35 | Homburg (CVT)<br>PI: Piergiorgio Lochner<br>Germany | CVT<br>N 6/5 | As for Homburg (row 8). | As for Homburg (row 8). |
| 36 | John Hunter Hospital, University of Newcastle, Newcastle<br>PI: Carlos Garcia-Esperon<br>Australia | CVT<br>N 4/3<br>CVT registry (ICVTC) | See ICVTC note below table. | The Hunter New England Research Ethics Committee approved a waiver of consent under the section 2.3.10 of the National Statement of Ethic Conduct in Human research, 2007 protocol. |
| 37 | South Limburg<br>PI: Rob Rouhl<br>Netherlands | ICH (excluded, 0 final)<br>N 138/0<br>ICH cohort submitted (R. Rouhl) but contributed 0 patients to the final analysis. | Excluded from analysis (0 eligible in final dataset). | Institutional approval. Source: questionnaire (South Limburg). |

**International CVT Consortium (ICVTC): collective methods (rows #4, 13, 14, 18 (the Sant Pau CVT cases), 23, 24, 27-30, 32, 34, 36).** CVT data were collected within the International Cerebral Venous Thrombosis Consortium, a multicenter international cohort of consecutive adults with CVT comprising 13 hospital-based registries from 12 countries (inclusion periods overall Dec 1994-Jun 2021), enrolled prospectively or retrospectively with routine follow-up (typically 6 and/or 12 months) and standardized case report forms capturing delayed seizures. Patients with epilepsy before CVT, follow-up <8 days, or missing late-seizure status were excluded; seizures were diagnosed by treating physicians per the ILAE definition. Each center obtained local ethics approval and written informed consent where required by national law. Source: International CVT Consortium (Lindgren, Coutinho, et al.).

**eTable 3.** *Sensitivity of the LHH-derived zone boundaries to alternative RRR and NNH*

| Moderate-risk entry, recurrence CIF (%) at LHH = 1.5 | NNH 10 | NNH 15 | NNH 19 | NNH 25 | NNH 30 |
|---|---|---|---|---|---|
| RRR 0.15 | 100.0 | 66.7 | 52.6 | 40.0 | 33.3 |
| RRR 0.20 | 75.0 | 50.0 | 39.5 | 30.0 | 25.0 |
| RRR 0.25 | 60.0 | 40.0 | 31.6 | 24.0 | 20.0 |
| RRR 0.30 | 50.0 | 33.3 | 26.3 | 20.0 | 16.7 |
| RRR 0.40 | 37.5 | 25.0 | 19.7 | 15.0 | 12.5 |

| LHH at the fixed high-risk boundary (ILAE 10-year recurrence CIF 60%) | NNH 10 | NNH 15 | NNH 19 | NNH 25 | NNH 30 |
|---|---|---|---|---|---|
| RRR 0.15 | 0.90 | 1.35 | 1.71 | 2.25 | 2.70 |
| RRR 0.20 | 1.20 | 1.80 | 2.28 | 3.00 | 3.60 |
| RRR 0.25 | 1.50 | 2.25 | 2.85 | 3.75 | 4.50 |
| RRR 0.30 | 1.80 | 2.70 | 3.42 | 4.50 | 5.40 |
| RRR 0.40 | 2.40 | 3.60 | 4.56 | 6.00 | 7.20 |

Upper panel: recurrence-CIF level of the lower zone boundary (LHH = 1.5) under the identity LHH = NNH x CIF x RRR, i.e. CIF = LHH / (NNH x RRR), capped at 100%. Lower panel: the upper zone boundary is fixed at the ILAE 60% 10-year recurrence threshold; the table gives the corresponding LHH = NNH x 0.60 x RRR. Base case (NNH 19, RRR 0.25): moderate-risk entry at CIF 31.6% and high-risk entry at CIF 60%, i.e. LHH 2.85. Lower RRR or lower NNH moves the lower boundary to a higher CIF and narrows the moderate zone; the moderate zone disappears (lower boundary at or above 60%) only when both are low (RRR 0.15 with NNH 10 or 15; RRR 0.20 with NNH 10; RRR 0.25 with NNH 10).

Abbreviations. CIF = cumulative incidence function; LHH = likelihood of being helped versus harmed.

**eTable 4.** *Inverse-probability-weighting covariate balance*

| Stroke subtype | Covariate | SMD before IPW | SMD after IPW |
|---|---|---|---|
| IS | Male sex | +0.088 | +0.004 |
| IS | Age (years) | -0.205 | -0.040 |
| IS | NIHSS 6-14 | +0.111 | +0.013 |
| IS | NIHSS ≥15 | +0.107 | +0.015 |
| IS | Large-artery atherosclerosis | +0.206 | +0.044 |
| IS | Cardioembolic mechanism | -0.083 | -0.017 |
| IS | Small-vessel disease | -0.132 | -0.038 |
| IS | MCA territory involvement | +0.151 | +0.025 |
| IS | Cortical involvement | +0.258 | +0.022 |
| IS | Reperfusion therapy | +0.007 | +0.014 |
| ICH | Male sex | -0.058 | +0.008 |
| ICH | Age (years) | -0.010 | -0.031 |
| ICH | NIHSS 6-14 | -0.062 | -0.028 |
| ICH | NIHSS ≥15 | -0.091 | -0.087 |
| ICH | Lobar hematoma | +0.356 | +0.061 |
| ICH | Hematoma volume >10 mL | -0.027 | -0.043 |
| ICH | Neurosurgical intervention | +0.216 | +0.115 |
| SAH | Male sex | +0.116 | +0.100 |
| SAH | Neurosurgical surgery | -0.181 | -0.054 |
| SAH | Early/acute-onset seizure | -0.494 | -0.358 |
| SAH | Age (years) | +0.048 | -0.076 |
| SAH | mRS at discharge | +0.372 | +0.123 |
| SAH | VASOGRADE | +0.289 | +0.169 |
| CVT | Male sex | +0.052 | +0.042 |
| CVT | Intracranial hemorrhage at presentation | -0.118 | -0.032 |
| CVT | Subdural hematoma | -0.150 | +0.007 |
| CVT | Decompressive surgery | +0.010 | +0.005 |
| CVT | Early/acute-onset seizure | +0.239 | +0.035 |
| CVT | Age (years) | -0.152 | -0.022 |

Standardized mean differences (SMD) before and after stabilized IPW on the final analysis cohort, for the covariates entered in each subtype-specific propensity model (treatment = any ASM). Age was entered as a continuous covariate. All post-IPW |SMD| were small (max 0.35, SAH early/acute-onset seizure), indicating adequate balance.

Abbreviations. ASM = antiseizure medication; CVT = cerebral venous thrombosis; ICH = intracerebral hemorrhage; IPW = inverse probability weighting; IS = ischemic stroke; mRS = modified Rankin Scale; NIHSS = National Institutes of Health Stroke Scale; SAH = subarachnoid hemorrhage.

**eTable 5.** *BIC-based search for a binary first-seizure timing cutoff*

| Analysis | Final N | Recurrences | BIC-optimal cutoff (days) | BIC at optimum | ΔBIC vs day 7 | ΔBIC vs day 30 | Plausible range ΔBIC ≤ 2 (days) | Plausible range ΔBIC ≤ 6 (days) | 60% upward crossing (days) | Later fall below 60% (days) |
|---|---|---|---|---|---|---|---|---|---|---|
| Pooled | 2212 | 630 | 20 | 7276.5 | −20.3 | −17.2 | 12-20 | 11-29 | - | - |
| IS | 931 | 277 | 20 | 3441.0 | −10.7 | −3.2 | 19-50 | 12-65 | 71 | 492 |
| ICH | 715 | 209 | 12 | 2411.6 | −9.6 | −11.3 | 12-29 | 11-29 | 41 | 692 |
| SAH | 338 | 101 | 1 | 1065.7 | −14.3 | −12.4 | 1 | 1 | 2 | 4 |
| CVT | 450 | 97 | 22 | 1009.1 | −14.0 | −9.3 | 11-28 | 11-29 | 9 | - |

Bayesian information criterion (BIC) search across candidate binary cut-points (days 1-30 and 35-90 in 5-day steps; both groups ≥20 patients and ≥5 recurrences) in stabilized-IPW Fine-Gray models on the final analysis cohort; BIC = −2 log-likelihood + k × log(number of recurrences). The pooled model is adjusted for sex, age ≥65 years and stroke severity and stratified by stroke subtype (n=2212 with complete covariates). The two right-hand columns give the zone boundaries of the canonical spline (same model as the risk zones; "Later fall below 60%" = first day after the high-risk zone) for comparison with the binary optimum.

Abbreviations. CVT = cerebral venous thrombosis; ICH = intracerebral hemorrhage; IPW = inverse probability weighting; IS = ischemic stroke; SAH = subarachnoid hemorrhage.

**eTable 6.** *Risk-zone difference and trajectory tests*

| **Within-subtype Gray K-sample (zones Low/Moderate/High)** | | | |
|---|---|---|---|
| Stroke subtype | $\chi^2$ | df | p |
| IS | 103.5 | 2 | <0.001 |
| ICH | 81.3 | 2 | <0.001 |
| SAH | 26.5 | 2 | <0.001 |
| CVT | 44.8 | 1 | <0.001 |

| **Between-subtype and trajectory tests** | **statistic** | **df** | **p** |
|---|---|---|---|
| Gray 4-sample (recurrence CIF, unadjusted) | 19.4 | 3 | <0.001 |
| Fine-Gray joint Wald on stroke subtype (adjusted for ns(log1p_day, df=3)) | 14.1 | 3 | 0.003 |
| C1 pseudo-value GAM (stroke subtype difference smooths; full vs null F-test) | 2.6 | 9.8 | 0.004 |
| C2 stabilized-IPW Fine-Gray robust Wald on stroke subtype x ns(log1p_day, df=3) interaction | 48.9 | 9 | <0.001 |

Analyses A (within-subtype), B (between-subtype) and C (trajectory). All on the final analysis cohort (n=2434); the four stroke subtypes (SAH as a single stratum) are compared on a common three-degree-of-freedom spline basis; C2 uses stabilized inverse-probability weights.

Abbreviations. CIF = cumulative incidence function; CVT = cerebral venous thrombosis; ICH = intracerebral hemorrhage; IPW = inverse probability weighting; IS = ischemic stroke; SAH = subarachnoid hemorrhage.

**eTable 7.** *Bootstrap cutoff stability and zone classification robustness*

| Stratum | Moderate-risk entry day (CIF 31.6%): point estimate; bootstrap median (95% PI; % of resamples) | High-risk zone (CIF ≥60%): entry day and last day, point estimate; bootstrap median (95% PI; % of resamples) |
|---|---|---|
| IS | 15; 15 (7-24; 100%) | entry 71; 70.5 (43-138; 99%); last day 491; 489 (302-746; 98%) |
| ICH | 9; 9 (4-17; 96%) | entry 41; 40 (24-77; 100%); last day 691; 626 (359-957; 85%) |
| SAH | 1; 1 (1-2; 71%) | entry 2; 2 (1-40; 97%); last day 3; 3 (2-198; 97%) |
| CVT | Moderate from day 0 (no low-risk zone) | entry 9; 9.5 (2-39; 100%); no return to moderate |

| Stroke subtype | Zone | 10-y CIF, point estimate (bootstrap 95% PI) | % sig / % monotone |
|---|---|---|---|
| IS | LOW | 25% (20-30) | 100% / 100% |
| IS | MED | 45% (35-53) | 100% / 100% |
| IS | HIGH | 67% (59-74) | 100% / 100% |
| ICH | LOW | 23% (18-30) | 100% / 99.9% |
| ICH | MED | 53% (44-62) | 100% / 99.9% |
| ICH | HIGH | 71% (61-80) | 100% / 99.9% |
| SAH | LOW | 22% (16-30) | 99.9% / 95.9% |
| SAH | MED | 44% (37-55) | 99.9% / 95.9% |
| SAH | HIGH | 69% (44-89) | 99.9% / 95.9% |
| CVT | MED | 38% (28-50) | 100% / 100% |
| CVT | HIGH | 92% (75-99) | 100% / 100% |

Upper panel: zone boundaries of the stabilized-IPW Fine-Gray spline (point estimate, as used for the risk zones) and of the same model refitted in 1000 bootstrap resamples (median, 95% percentile interval, and percentage of resamples in which the boundary exists). Lower panel: zones held fixed; 10-year recurrence cumulative incidence point estimate (as in the main text and eTable 11) with the 95% percentile interval of 1000 bootstrap resamples, and the percentage of resamples in which the zones differed significantly (Wald $p<0.05$) and were monotonically ordered. Intervals in this table are bootstrap percentile intervals; the main text reports model-based 95% CIs for the same point estimates. Bootstrap medians of the zone entry days lie within 1 day of the point estimates for all 4 stroke subtypes, whereas the medians of the days on which the curves leave the high-risk zone are less stable (ischemic stroke, 489 vs 491; intracerebral hemorrhage, 626 vs 691).

Abbreviations. CIF = cumulative incidence function; CVT = cerebral venous thrombosis; ICH = intracerebral hemorrhage; IPW = inverse probability weighting; IS = ischemic stroke; PI = prediction interval; SAH = subarachnoid hemorrhage.

**eTable 8.** *Follow-up / cohort sensitivity (leave-one-cohort-out)*

| Stroke subtype | Full-cohort high-vs-low 10-y CIF gap (Gray p) | Leave-one-cohort-out gap range (n cohorts) | Zone Gray test across LOO |
|---|---|---|---|
| IS | 42 pp (<0.001) | 39-44 pp (14) | all p<0.05 |
| ICH | 47 pp (<0.001) | 6-56 pp (6) | all p<0.05 |
| SAH | 47 pp (<0.001) | 44-58 pp (3) | all p<0.05 |
| CVT | 54 pp (<0.001) | 43-58 pp (6) | all p<0.05 |

Gap = 10-year recurrence cumulative incidence of the high-risk minus the low-risk zone (moderate-risk zone for CVT, which has no low-risk zone), from the stabilized-IPW Fine-Gray zone model used in the main text, with weights re-estimated after each omission; recomputed after dropping each cohort contributing ≥25 individuals to that subtype. The zone difference was tested with Gray's K-sample test. pp = percentage points. The 29 retained subtype-cohort units cover 2184 of 2434 individuals (90%); per-center counts are in eTable 1. Smaller contributors were retained in all models but not removed individually. The intracerebral hemorrhage gap range is the widest, narrowing to 6 percentage points in one omission while remaining statistically significant.

Abbreviations. CIF = cumulative incidence function; CVT = cerebral venous thrombosis; ICH = intracerebral hemorrhage; IPW = inverse probability weighting; IS = ischemic stroke; pp = percentage points; SAH = subarachnoid hemorrhage.

**eTable 9.** *Screening and eligibility cascade by stroke subtype*

| Stroke subtype | Screened | Missing follow-up | Missing recurrence | Missing treatment | Missing seizure timing | <30 d observation | Eligible |
|---|---|---|---|---|---|---|---|
| IS | 1106 | 88 | 0 | 0 | 69 | 18 | 931 |
| ICH | 956 | 219 | 0 | 0 | 3 | 19 | 715 |
| SAH | 390 | 9 | 0 | 0 | 2 | 41 | 338 |
| CVT | 526 | 3 | 0 | 0 | 5 | 68 | 450 |
| Total | 2978 | 319 | 0 | 0 | 79 | 146 | 2434 |

Hierarchical, mutually-exclusive exclusion reasons; counts sum to screened minus eligible. CVT first-seizure days were pooled to day 0 before the >=30-day observation rule.

Abbreviations. CVT = cerebral venous thrombosis; ICH = intracerebral hemorrhage; IS = ischemic stroke; SAH = subarachnoid hemorrhage.

**eTable 10.** *Potential overdiagnosis of epilepsy under the current 7-day cutoff (ischemic stroke and intracerebral hemorrhage).*

| **Stroke subtype** | **First seizures after day 7 (remote), n** | **Reaching ≥60% high-risk zone, n** | **Below 60% (potential overdiagnosis), n (%)** |
|---|---|---|---|
| Ischemic stroke | 464 | 255 | 209 (45%) |
| Intracerebral hemorrhage | 336 | 210 | 126 (38%) |
| Combined | 800 | 465 | 335 (42%) |

Under the current definition, a first seizure occurring after day 7 is classified as remote symptomatic (unprovoked) and, in a patient with a structural brain lesion, may prompt a diagnosis of epilepsy and long-term antiseizure medication. Later-onset (day >7) counts are from Table 1. A first seizure was counted as reaching the epilepsy-level threshold when its day fell in the ≥60% 10-year-recurrence (high-risk) zone, ischemic stroke, days 71-491; intracerebral hemorrhage, days 41-691 (eMethods and eTable 11); in both stroke subtypes every high-risk-zone first seizure occurs after day 7. Percentages are of later-onset seizures. Of the first seizures below 60%, 132 (IS) and 74 (ICH) occurred after the end of the high-risk zone (day ≥492 and ≥692, respectively), where estimates rest on fewer patients with long follow-up. Subarachnoid hemorrhage and cerebral venous thrombosis are omitted because their high-risk-zone estimates rest on small numbers.

**eTable 11.** *Cumulative incidence of seizure recurrence by risk zone at 1, 2, 3, 5 and 10 years*

| Stroke subtype, risk zone (n) | 1 year | 2 years | 3 years | 5 years | 10 years |
|---|---|---|---|---|---|
| IS, Low (n=487) | 15 (12-18) | 19 (16-23) | 21 (17-25) | 23 (19-27) | 25 (20-30) |
| IS, Moderate (n=189) | 28 (22-35) | 36 (29-44) | 38 (31-47) | 42 (34-50) | 45 (36-54) |
| IS, High (n=255) | 46 (40-53) | 56 (49-63) | 59 (53-66) | 63 (56-70) | 67 (60-74) |
| ICH, Low (n=256) | 13 (10-17) | 16 (13-21) | 18 (13-23) | 20 (16-26) | 23 (18-30) |
| ICH, Moderate (n=249) | 33 (28-40) | 40 (34-48) | 43 (36-50) | 48 (40-56) | 53 (44-62) |
| ICH, High (n=210) | 48 (40-57) | 57 (48-65) | 59 (51-68) | 65 (56-73) | 71 (61-79) |
| SAH, Low (n=168) | 9 (6-14) | 12 (8-18) | 15 (10-21) | 19 (13-26) | 22 (16-30) |
| SAH, Moderate (n=144) | 20 (15-27) | 26 (20-34) | 31 (24-39) | 38 (31-47) | 44 (36-53) |
| SAH, High (n=26) | 37 (23-57) | 46 (29-68) | 53 (35-75) | 63 (42-83) | 69 (48-89) |
| CVT, Moderate (n=414) | 13 (10-17) | 19 (15-24) | 22 (17-28) | 23 (18-29) | 38 (29-49) |
| CVT, High (n=36) | 53 (37-70) | 67 (50-83) | 74 (56-88) | 75 (58-90) | 92 (77-99) |

Abbreviations. CVT = cerebral venous thrombosis; ICH = intracerebral hemorrhage; IS = ischemic stroke; SAH = subarachnoid hemorrhage. Values are the cumulative incidence of seizure recurrence (%) with 95% CI, estimated from competing-risk (Fine-Gray) models with stabilized inverse-probability-of-treatment weighting and all-cause death as the competing event.

### eFigures

**eFigure 1.** *Patient flow, pooled cohort and the four subtype-specific subcohorts.*

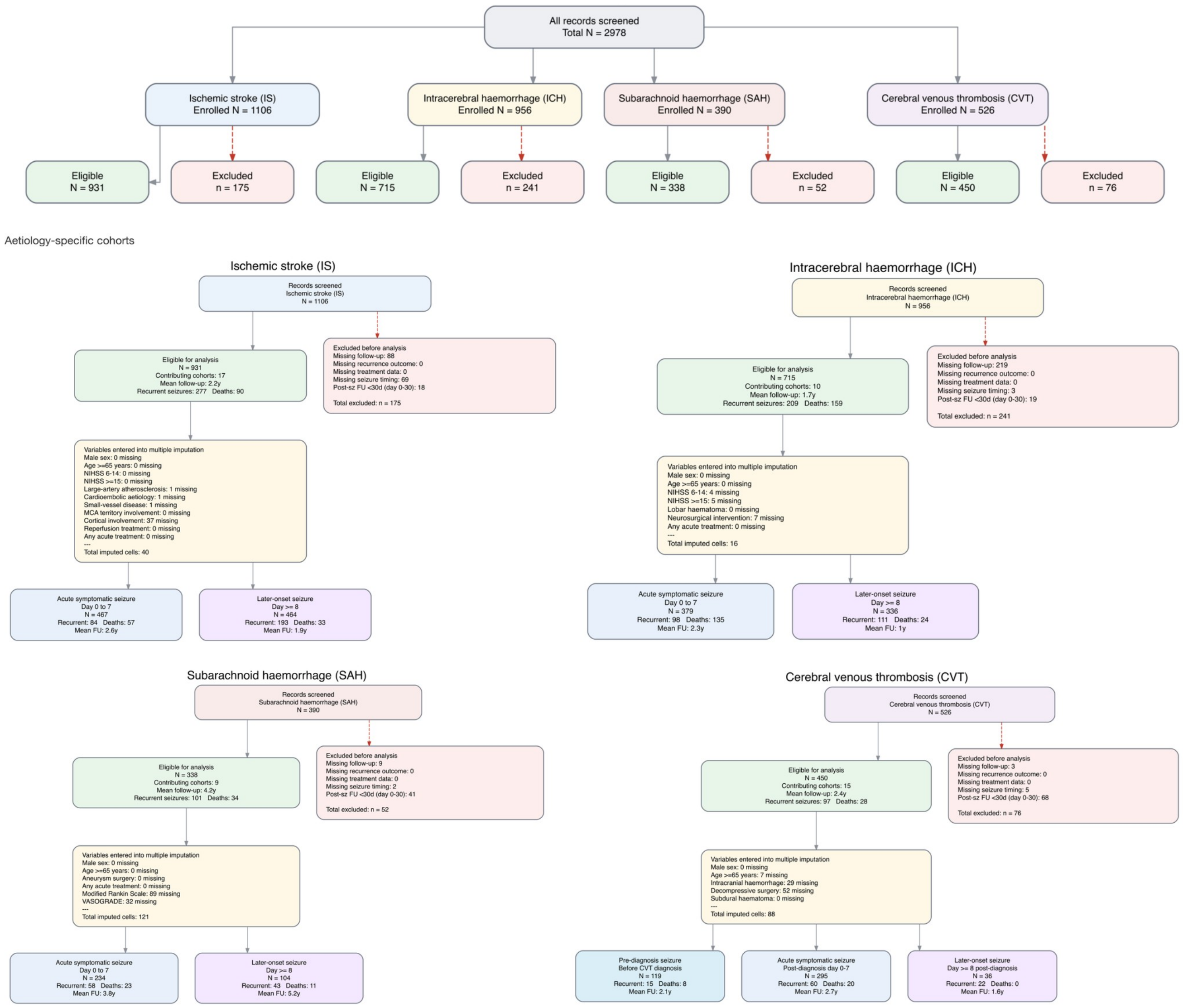

Patient flow for the pooled cohort and the four subtype-specific subcohorts. The top panel shows the pooled, CONSORT-style cascade from enrollment to the final analysis set. The lower four panels show the subtype-specific cascades for ischemic stroke (IS), intracerebral hemorrhage (ICH), subarachnoid hemorrhage (SAH) and cerebral venous thrombosis (CVT), with exclusion reasons applied hierarchically: pre-existing epilepsy or a seizure judged unrelated to the stroke; missing or internally inconsistent first-seizure timing, follow-up or outcome data; and fewer than 30 days of observable follow-up from the index event. Each cascade also summarizes the imputation applied to the subtype-specific model covariates and the resulting first-seizure timing groups (acute, days 0-7; later, day ≥8; and, for CVT, pre-diagnosis seizures pooled to day 0). The final analysis cohort comprised 2434 patients (IS 931, ICH 715, SAH 338, CVT 450), with 684 seizure recurrences and 311 competing deaths.

Abbreviations. AJ = Aalen-Johansen; ASM = antiseizure medication; CI = confidence interval; CIF = cumulative incidence function; CVT = cerebral venous thrombosis; ESS = effective sample size; ICH = intracerebral hemorrhage; IPW = inverse probability weighting; IS = ischemic stroke; LHH = likelihood of being helped versus harmed; mRS = modified Rankin Scale; NIHSS = National Institutes of Health Stroke Scale; PI = prediction interval; pp = percentage points; SAH = subarachnoid hemorrhage.

**eFigure 2.** *Zone-level Aalen-Johansen CIF and reliability diagnostics.*

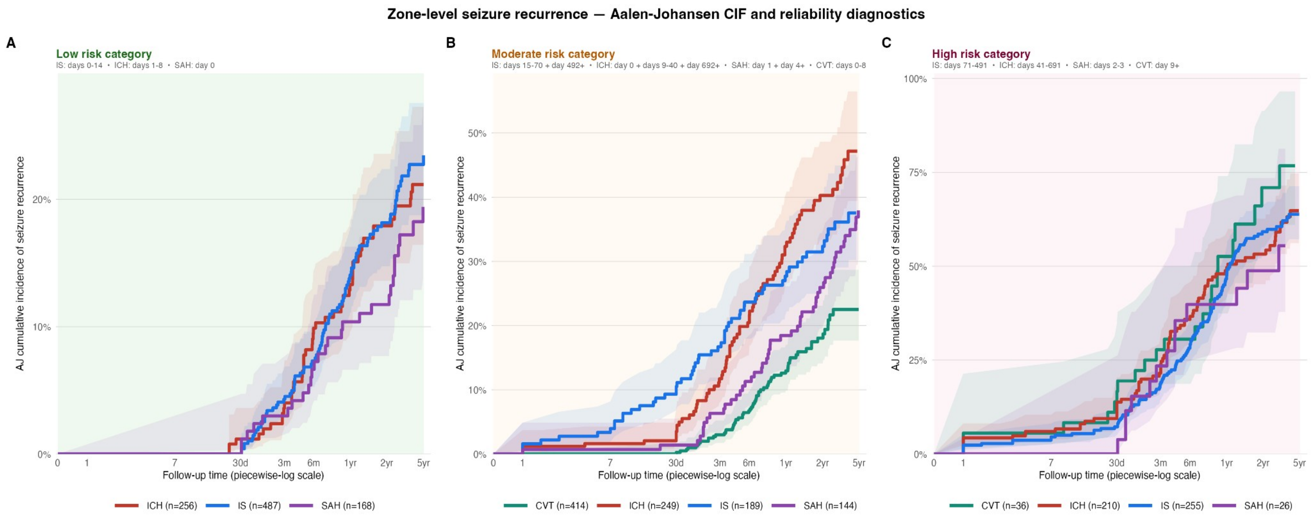

D

Reliable (ESS > 50%) | Borderline (25–50%) | Unstable (15–25%) | Critical (< 15%)

| | n | ESS (n / %) | | n at risk (1yr / 2yr / 5yr) | | | 95% CI ± pp (1yr / 2yr / 5yr) | | |
|---|---|---|---|---|---|---|---|---|---|
| IS | 255 | 252 | 99% | 95 | 58 | 25 | ±6.9 | ±6.9 | ±7.1 |
| ICH | 210 | 205 | 97% | 53 | 37 | 11 | ±8.6 | ±8.7 | ±9.3 |
| SAH | 26 | 26 | 100% | 14 | 8 | 3 | ±20.0 | ±20.6 | ±21.7 |
| CVT | 36 | 34 | 94% | 12 | 6 | 1 | ±18.3 | ±18.1 | ±17.8 |
| IS | 189 | 187 | 99% | 94 | 70 | 30 | ±7.0 | ±7.5 | ±8.2 |
| ICH | 249 | 244 | 98% | 92 | 61 | 23 | ±7.1 | ±7.6 | ±8.5 |
| SAH | 144 | 143 | 99% | 106 | 88 | 53 | ±6.5 | ±7.4 | ±8.5 |
| CVT | 414 | 404 | 98% | 245 | 120 | 45 | ±3.6 | ±4.5 | ±5.5 |
| IS | 487 | 479 | 98% | 301 | 226 | 87 | ±3.3 | ±3.8 | ±4.6 |
| ICH | 256 | 254 | 99% | 123 | 91 | 46 | ±4.4 | ±5.0 | ±5.4 |
| SAH | 168 | 164 | 98% | 133 | 112 | 60 | ±4.8 | ±5.1 | ±6.8 |
| CVT | 0 | – | – | – | – | – | – | – | – |

Unadjusted (Aalen-Johansen) cumulative incidence of seizure recurrence by risk zone, with reliability diagnostics. Panels A, B and C show the Aalen-Johansen cumulative incidence of recurrence (step curves with 95% CI) for the Low, Moderate and High risk zones, respectively, with the four stroke subtypes (IS, ICH, SAH, CVT) overlaid within each panel. These unadjusted estimates complement the stabilized-IPW Fine-Gray zone curves in Figures 2 and 3 by showing the raw recurrence experience underlying each zone. Panel D is a reliability heatmap of every zone × stroke subtype cell, rated on a four-tier scale (reliable, borderline, unstable, critical) that combines effective sample size as a fraction of zone N, the number at risk, and the width of the 10-year confidence interval; the tiers flag cells whose estimates rest on limited statistical support, notably the small SAH and CVT high-risk zones.

Abbreviations. AJ = Aalen-Johansen; ASM = antiseizure medication; CI = confidence interval; CIF = cumulative incidence function; CVT = cerebral venous thrombosis; ESS = effective sample size; ICH = intracerebral hemorrhage; IPW = inverse probability weighting; IS = ischemic stroke; LHH = likelihood of being helped versus harmed; mRS = modified Rankin Scale; NIHSS = National Institutes of Health Stroke Scale; PI = prediction interval; pp = percentage points; SAH = subarachnoid hemorrhage.

**eFigure 3.** *Bootstrap stability of the risk-zone cutoffs.*

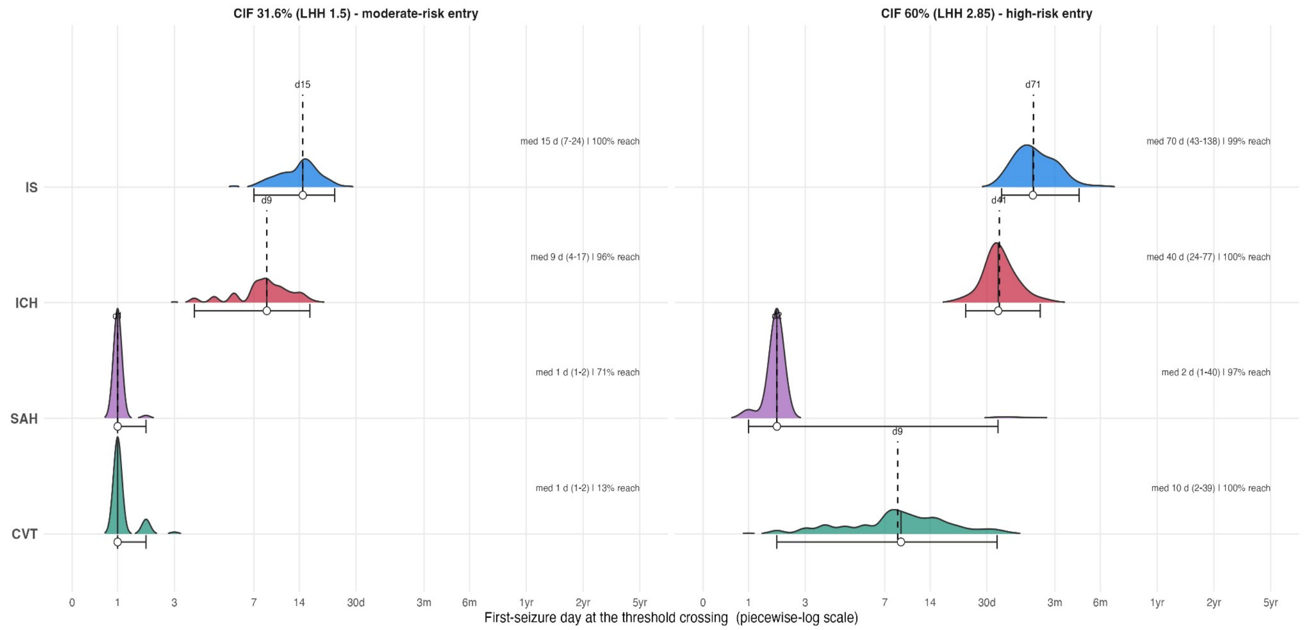

Sample-to-sample stability of the risk-zone cutoff days. For each stratum (IS, ICH, SAH, and CVT) the spline risk curve was re-estimated in 1000 stabilized-IPW Fine-Gray bootstrap resamples (window-stratified resampling for the day-0-dominated SAH and CVT strata), and the zone boundaries were re-derived with the same rule as the main analysis: entry into the moderate zone (10-year recurrence cumulative incidence 31.6%, LHH 1.5) and into the high-risk zone (60%, LHH 2.85). The ridge (density) plots show the bootstrap distribution of these crossing days, annotated with the bootstrap median, the 95% percentile interval, and the point-estimate zone-entry day used in the main analysis (dashed line). The spread of each distribution, and the proportion of resamples in which the curve never reaches 60%, indicate how sample-dependent the exact boundary day is.

Abbreviations. AJ = Aalen-Johansen; ASM = antiseizure medication; CI = confidence interval; CIF = cumulative incidence function; CVT = cerebral venous thrombosis; ESS = effective sample size; ICH = intracerebral hemorrhage; IPW = inverse probability weighting; IS = ischemic stroke; LHH = likelihood of being helped versus harmed; mRS = modified Rankin Scale; NIHSS = National Institutes of Health Stroke Scale; PI = prediction interval; pp = percentage points; SAH = subarachnoid hemorrhage.

**eFigure 4.** *Robustness of the published risk-zone classification (fixed cutoffs, bootstrapped).*

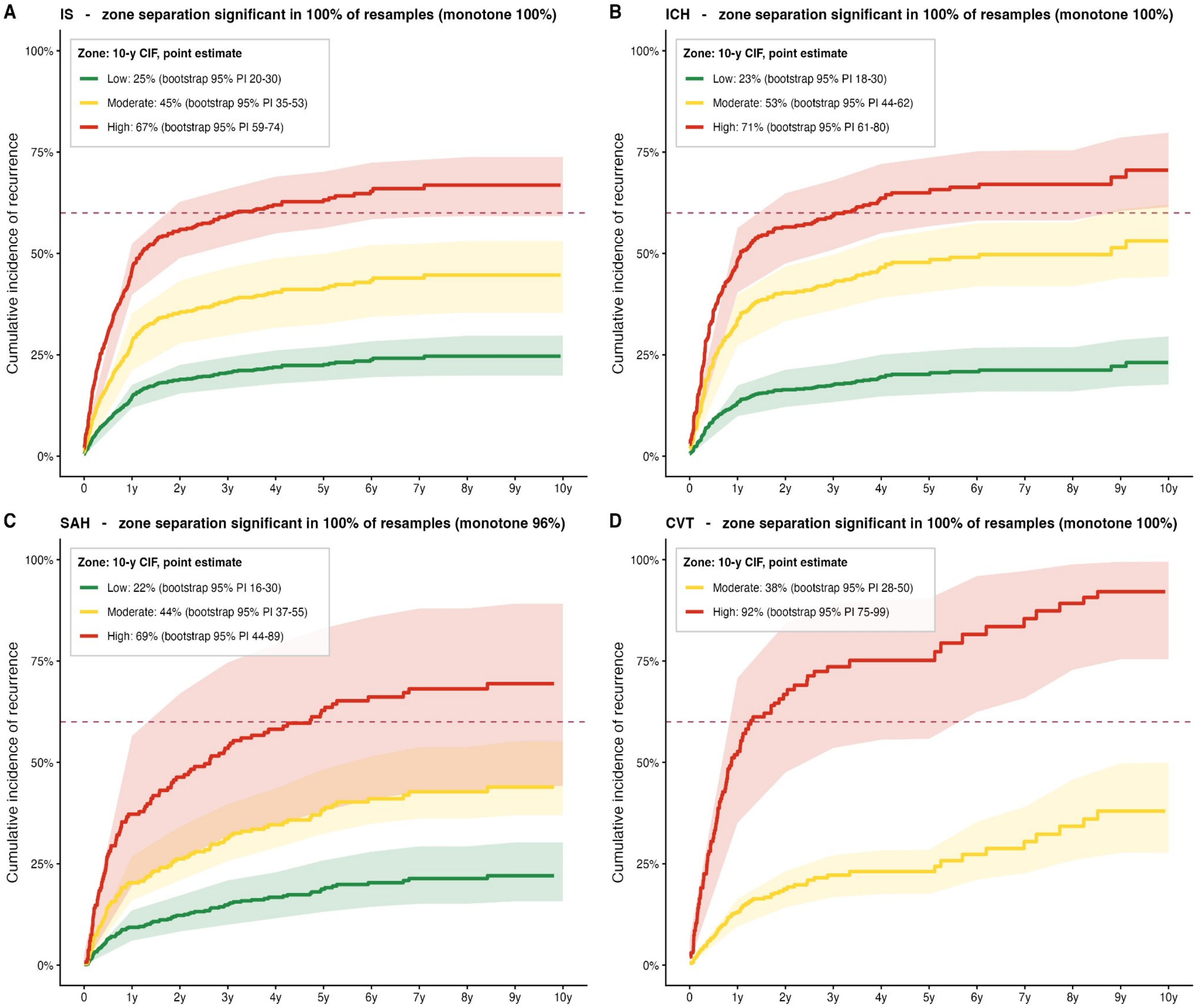

Robustness of the published risk-zone classification when the cutoffs are held fixed. With the zone boundaries fixed at their published values, the stabilized-IPW Fine-Gray within-zone curves were recomputed across 1000 bootstrap resamples. Each panel shows, by stroke subtype, the within-zone recurrence cumulative incidence (lines: point estimates, as in Figures 2 and 3) with the 95% percentile interval of the bootstrap resamples (ribbons); the panel title gives the proportion of resamples in which the zones remained significantly distinct (Wald test of the zone term) and correctly ordered (monotone). The zones separated significantly in 99.9-100% of resamples and remained monotone in 100% (IS), 99.9% (ICH), 100% (CVT) and 96% (SAH) of resamples; the SAH high-risk zone (days 2-3) is the least precise owing to small numbers.

Abbreviations. AJ = Aalen-Johansen; ASM = antiseizure medication; CI = confidence interval; CIF = cumulative incidence function; CVT = cerebral venous thrombosis; ESS = effective sample size; ICH = intracerebral hemorrhage; IPW = inverse probability weighting; IS = ischemic stroke; LHH = likelihood of being helped versus harmed; mRS = modified Rankin Scale; NIHSS = National Institutes of Health Stroke Scale; PI = prediction interval; pp = percentage points; SAH = subarachnoid hemorrhage.

**eFigure 5.** *All-cause mortality after a first poststroke seizure, IS and ICH.*

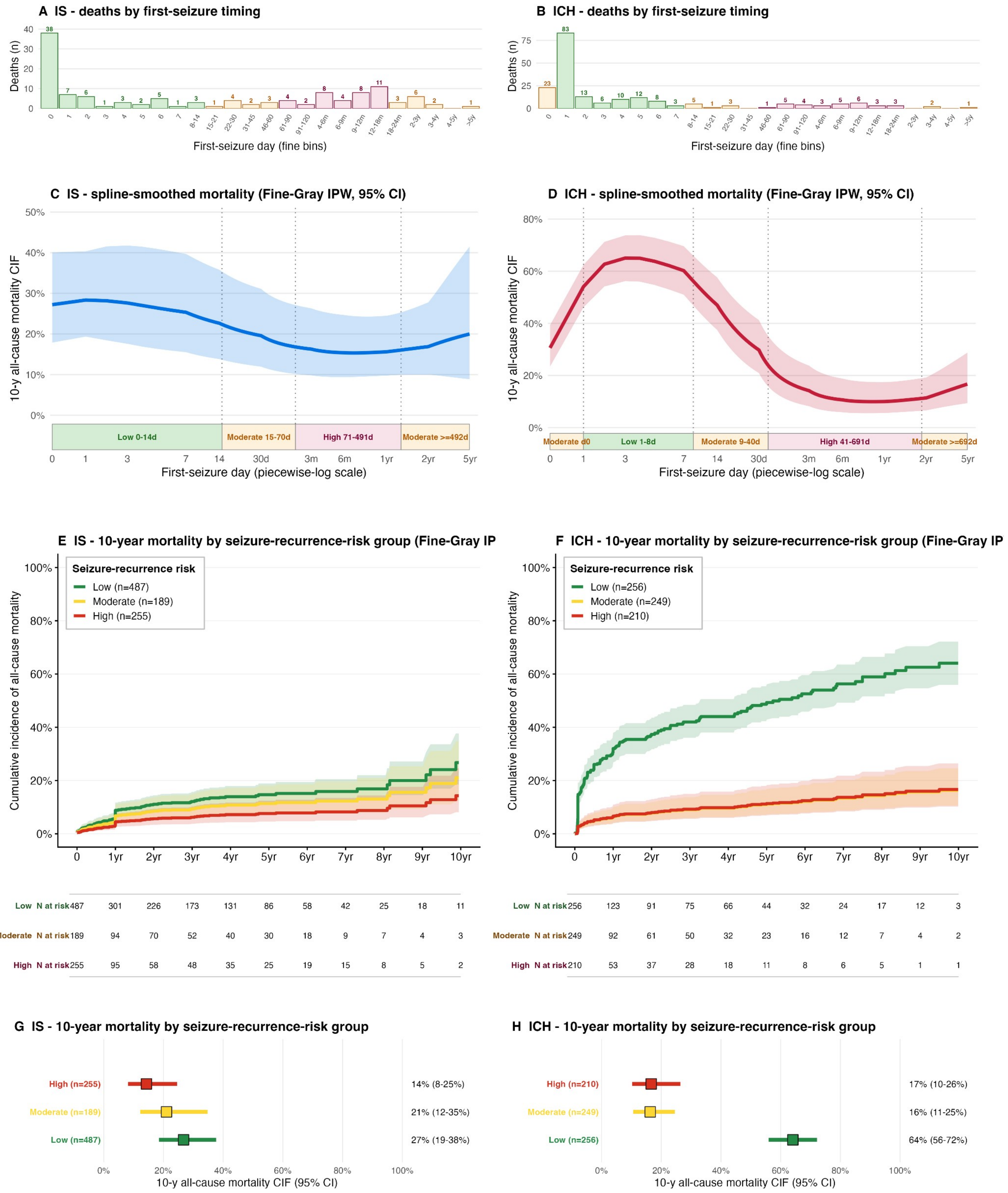

All-cause mortality after a first poststroke seizure in ischemic stroke (IS, left column) and intracerebral hemorrhage (ICH, right column), using the same panel template as Figure 2 but with all-cause death without prior recurrence as the event and seizure recurrence as the competing risk. Panels A-B show the distribution of first-seizure timing (histograms). Panels C-D show the IPW Fine-Gray spline-smoothed 10-year mortality cumulative incidence (solid line, shaded 95% CI), overlaid on the same recurrence-risk zones as Figure 2. Panels E-F show the 10-year mortality cumulative incidence within each seizure-recurrence-risk group (step curves with numbers at risk), and panels G-H the corresponding per-group mortality forest plots. Mortality was highest in the low-recurrence first-seizure groups.

Abbreviations. AJ = Aalen-Johansen; ASM = antiseizure medication; CI = confidence interval; CIF = cumulative incidence function; CVT = cerebral venous thrombosis; ESS = effective sample size; ICH = intracerebral hemorrhage; IPW = inverse probability weighting; IS =

ischemic stroke; LHH = likelihood of being helped versus harmed; mRS = modified Rankin Scale; NIHSS = National Institutes of Health Stroke Scale; PI = prediction interval; pp = percentage points; SAH = subarachnoid hemorrhage.

**eFigure 6.** *All-cause mortality after a first poststroke seizure, SAH and CVT.*

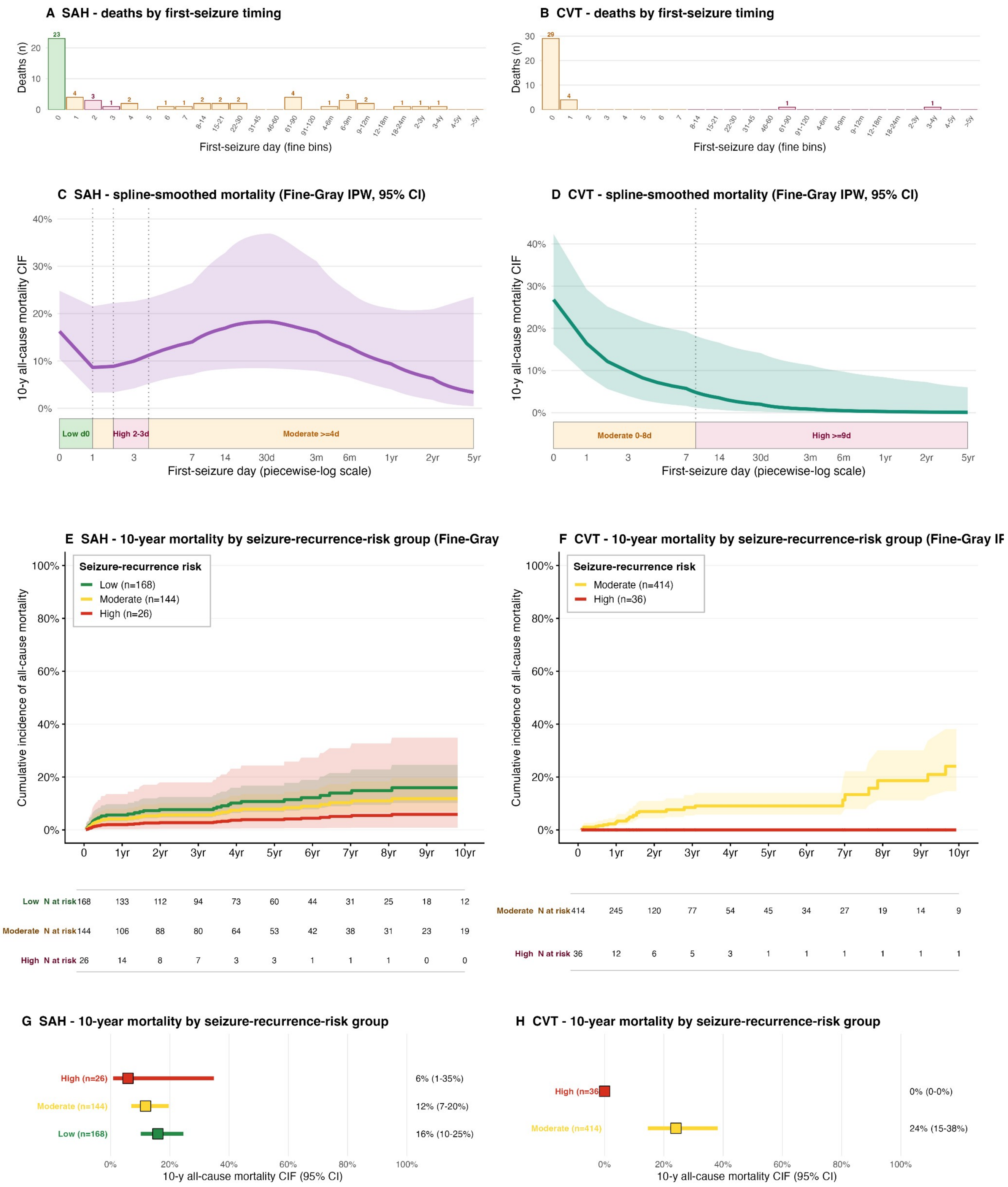

| | 0 | 1yr | 2yr | 3yr | 4yr | 5yr | 6yr | 7yr | 8yr | 9yr | 10yr |
|---|---|---|---|---|---|---|---|---|---|---|---|
| SAH Low N at risk | 168 | 133 | 112 | 94 | 73 | 60 | 44 | 31 | 25 | 18 | 12 |
| SAH Moderate N at risk | 144 | 106 | 88 | 80 | 64 | 53 | 42 | 38 | 31 | 23 | 19 |
| SAH High N at risk | 26 | 14 | 8 | 7 | 3 | 3 | 1 | 1 | 1 | 0 | 0 |
| CVT Moderate N at risk | 414 | 245 | 120 | 77 | 54 | 45 | 34 | 27 | 19 | 14 | 9 |
| CVT High N at risk | 36 | 12 | 6 | 5 | 3 | 1 | 1 | 1 | 1 | 1 | 1 |

All-cause mortality after a first poststroke seizure in subarachnoid hemorrhage (SAH, left column) and cerebral venous thrombosis (CVT, right column), constructed exactly as eFigure 5 (all-cause death without prior recurrence as the event, seizure recurrence as the competing risk; CVT pre-index seizures pooled to day 0). Panels A-B show first-seizure timing histograms; panels C-D the IPW Fine-Gray spline-smoothed 10-year mortality cumulative incidence (95% CI) over the recurrence-risk zones; panels E-F the 10-year mortality cumulative incidence by recurrence-risk group with numbers at risk; and panels G-H the per-group mortality forest plots. CVT has no low-recurrence-risk group, and the small CVT high-risk group had no deaths without prior recurrence; SAH and CVT later-window estimates rest on small numbers and should be interpreted as exploratory.

Abbreviations. AJ = Aalen-Johansen; ASM = antiseizure medication; CI = confidence interval; CIF = cumulative incidence function; CVT = cerebral venous thrombosis; ESS = effective sample size; ICH = intracerebral hemorrhage; IPW = inverse probability weighting; IS = ischemic stroke; LHH = likelihood of being helped versus harmed; mRS = modified Rankin Scale; NIHSS = National Institutes of Health Stroke Scale; PI = prediction interval; pp = percentage points; SAH = subarachnoid hemorrhage.
