## Supplement 2: Nonauthor Collaborators for "Timing of First Poststroke Seizure and Risk of Seizure Recurrence by Stroke Subtype"

The following investigators contributed data and/or scientific expertise to this study as nonauthor collaborators of the SeLECT-RS Study Group. Their affiliations, ORCID identifiers, and disclosures are listed in the eTable.

**SeLECT Consortium:** Giada Giovannini; Estevo Santamarina; José Álvarez-Sabín; Teresa Pinho e Melo; Marta Olivé; Frauke Roell; Zafeirenia Vlakou; Anastasios Bonakis; Georgios Tsivgoulis; Nico Deligas; Barbara Erdélyi-Canavese; Dominik Zieglgänsberger; Ansgar Felbecker; Philip Siebel; Gian Luigi Gigli; Annacarmen Nilo; Francesco Janes; Alessandra Burini; Domenico Maisano; Ana Rita Peralta; Michael Winklehner; Tim J von Oertzen; Judith N. Wagner; Vasiliki Poulidou; Martha Spilioti; Thomas Tegos; Atte Meretoja; Bernd Vorderwülbecke; Robert Terziev; Martin Holtkamp; Chiara Tatillo; Gilles Naeije; Julian Conrad; Stefan Evers; Andreas R. Luft; Miranda Stattmann; Anton Schmick; Maria Pagiantza; Giulio Bicciato; Lucia Sinka; Adam Strzelczyk; Carolina Ferreira-Atuesta; Mira Katan; Arianna Scartezzini; Federico Mazzacane; Francesco Valletta; Hui Jan Tan; Markus Arnold; Michael Knoflach; Patrick Kwan; Zhen Li.

**International CVT Consortium:** Turgut Tatlisumak; Katarina Jood; Marcel Arnold; José M Ferro; Patrícia Canhão; Antonio Arauz; Masoud Ghiasian; Maryam Mansour; Daniel Guisado-Alonso; Esme Ekizoglu; Nilüfer Yesilot; Mine Sezgin; Miguel A Barboza; Valentina Arnao; Paolo Aridon; Carlos Garcia-Esperon; João Morgadinho; Miguel Rodrigues.

**MUCH-Italy Investigators:** Anna Cavallini; Cristiano Azzini; Elisa Giorli; Giacomo Giacalone; Giorgio Silvestrelli; Manuel Cappellari; Marialuisa Zedde; Massimo Del Sette; Maurizio Paciaroni; Mauro Magoni; Rocco Salvatore Calabrò; Rossana Tassi; Simona Marcheselli; Simone Vidale.

**ILAE Acute Symptomatic Seizures Task Force:** Cecil D Hahn; Clio Aracelli Rubinos; Dong Zhou; Gagandeep Singh; Jo Wilmshurst; Johan Zelano; Lawrence J. Hirsch; Luis Otavio Caboclo; Melody T Asukile; Nick Abend; Nicola Specchio; Nirmeen A. Kishk; Ruta Yardi; Sahar Zafar; Simona Lattanzi; Stephane Auvin; Susanne Knake; Yamile Calle-Lopez.

**eTable. Nonauthor collaborators of the SeLECT-RS Study Group, with network attribution, affiliations, ORCID identifiers, and disclosures**

| **Name and network** | **Affiliation** | **ORCID** | **Disclosures** |
|---|---|---|---|
| Giada Giovannini, MD PhD *SeLECT Consortium* | Neurophysiology Unit and Epilepsy Centre, Neuroscience Department, Modena AOU, Modena, Italy. | 0000-0002-3585-5872 | Nothing to disclose. |
| Estevo Santamarina, MD PhD *SeLECT Consortium* | Epilepsy Unit, Department of Neurology, Vall d'Hebron Hospital Universitari, Barcelona; Universitat Autònoma de Barcelona, Bellaterra, Spain | 0009-0006-3940-8953 | ES reports research funding and speaker fees from, and paid consultancy for, UCB Pharma, Bial Pharmaceutical, Eisai, Arvelle, Angelini Pharma, Neuraxpharm, and Laboratorios Esteve. |
| José Álvarez-Sabín, PhD *SeLECT Consortium* | Epilepsy Unit, Department of Neurology, Vall d'Hebron Hospital Universitari, Barcelona; Universitat Autònoma de Barcelona, Bellaterra, Spain. | 0000-0002-8054-9209 | Nothing reported. |
| Teresa Pinho e Melo, MD *SeLECT Consortium* | Epilepsy Unit, Department of Neurology, Vall d'Hebron Hospital Universitari, Barcelona; Universitat Autònoma de Barcelona, Bellaterra, Spain. | | Nothing reported. |
| Marta Olivé *SeLECT Consortium* | Epilepsy Unit, Department of Neurology, Vall d'Hebron Hospital Universitari, Barcelona; Universitat Autònoma de Barcelona, Bellaterra, Spain. | 0000-0003-4637-1598 | Nothing reported. |
| Frauke Roell, MD *SeLECT Consortium* | Department of Neurology, Saarland University Medical Center, Homburg, Germany. | | Nothing reported. |

| Name and network | Affiliation | ORCID | Disclosures |
|---|---|---|---|
| Zafeirenia Vlakou, MD *SeLECT Consortium* | Second Department of Neurology, “Attikon” University Hospital, School of Medicine, National & Kapodistrian University of Athens, Athens, Greece | | Nothing to disclose. |
| Anastasios Bonakis, MD PhD *SeLECT Consortium* | Second Department of Neurology, Attikon University Hospital, National and Kapodistrian University of Athens, Athens, Greece. | | Nothing reported. |
| Georgios Tsivgoulis, MD PhD *SeLECT Consortium* | Second Department of Neurology, Attikon University Hospital, National and Kapodistrian University of Athens, Athens, Greece. | 0000-0002-0640-3797 | Nothing reported. |
| Nico Deligas, MD *SeLECT Consortium* | Department of Neurology, Kantonsspital St. Gallen, St Gallen, Switzerland | 0000-0001-9795-9127 | Nothing to disclose. |
| Barbara Erdélyi-Canavese, MD *SeLECT Consortium* | Department of Neurology, Kantonsspital St. Gallen, St Gallen, Switzerland. | | Nothing to disclose. |
| Dominik Zieglgänsberger, MD *SeLECT Consortium* | Department of Neurology, Cantonal Hospital, St. Gallen, Switzerland | 0009-0007-7112-304X | Nothing to disclose. |
| Ansgar Felbecker, MD *SeLECT Consortium* | University Hospital Bern, Department of Neurology, Bern, Switzerland | 0000-0001-9073-6617 | Nothing to disclose. |
| Philip Siebel, MD *SeLECT Consortium* | Neurologie am Rosenberg, Frauenfeld, Switzerland | 0009-0008-0827-0589 | Nothing to disclose. |
| Gian Luigi Gigli, MD *SeLECT Consortium* | Department of Medicine, University of Udine and Clinical Neurology, Udine University Hospital, Udine, Italy. | 0000-0003-3526-1414 | Nothing reported. |
| Annacarmen Nilo, MD *SeLECT Consortium* | Department of Medicine, University of Udine and Clinical Neurology, Udine University Hospital, Udine, Italy. | 0000-0001-9827-8059 | Nothing reported. |
| Francesco Janes, MD, PhD *SeLECT Consortium* | Clinic of Neurology, Head and Neck Department, Udine Academic Hospital | 0000-0003-1534-1409 | Nothing to disclose. |
| Alessandra Burini, MD *SeLECT Consortium* | Clinical Neurology, Department of Medicine (DMED), University of Udine, Udine, Italy Epilepsy Unit, Fondazione IRCCS Istituto Neurologico Carlo Besta, Milan, Italy | 0000-0002-8719-2441 | Nothing to disclose. |
| Domenico Maisano, MD *SeLECT Consortium* | Clinical Neurology, DMED, University of Udine, Udine, Italy. | | Nothing to disclose. |
| Ana Rita Peralta, MD *SeLECT Consortium* | Reference Centre for Refractory Epilepsy (member ERN EpiCARE), Hospital de Santa Maria-ULSSM, Lisbon, Portugal; Department of Neurology, Hospital de Santa Maria-ULSSM, Lisbon, Portugal; MUHCRI, Faculdade de Medicina, Universidade de Lisboa, Lisbon, Portugal. | 0000-0001-9449-431X | Nothing reported. |
| Michael Winklehner, MD *SeLECT Consortium* | Department of Neurology, Johannes Kepler University Linz, Kepler University Hospital, Linz, Austria; Clinical Research Institute for Neurosciences, Johannes Kepler University Linz and Kepler University Hospital, Linz, Austria | 0000-0003-4938-7827 | Nothing to disclose. |
| Tim J von Oertzen, MD FRCP *SeLECT Consortium* | Department of Neurology, Kepler University Hospital, Johannes Kepler University Linz, Linz, Austria. | 0000-0003-2164-7842 | Nothing reported. |

| Name and network | Affiliation | ORCID | Disclosures |
|---|---|---|---|
| Judith N. Wagner, MD *SeLECT Consortium* | Faculty of Medicine, University Duisburg-Essen, Essen, Germany | 0000-0002-0776-6821 | Nothing to disclose. |
| Vasiliki Poulidou, MD *SeLECT Consortium* | First Department of Neurology, AHEPA University Hospital, Aristotle University of Thessaloniki, Thessaloniki, Greece. | 0000-0002-0147-3948 | Nothing to disclose. |
| Martha Spilioti, MD *SeLECT Consortium* | First Department of Neurology, AHEPA University Hospital, Aristotle University of Thessaloniki, Thessaloniki, Greece. | | Nothing reported. |
| Thomas Tegos, MD *SeLECT Consortium* | First Department of Neurology, AHEPA University Hospital, Aristotle University of Thessaloniki, Thessaloniki, Greece. | | Nothing reported. |
| Atte Meretoja *SeLECT Consortium* | Department of Neurology, Helsinki University Hospital and University of Helsinki, Helsinki, Finland. | | Nothing reported. |
| Bernd Vorderwülbecke *SeLECT Consortium* | Epilepsy-Center Berlin-Brandenburg, Department of Neurology with Experimental Neurology, Charité – Universitätsmedizin Berlin, Berlin, Germany. | 0000-0002-0528-4001 | Nothing reported. |
| Robert Terziev, MD *SeLECT Consortium* | Department of Neurology, Charité - Universitätsmedizin Berlin, Berlin, Germany Berlin Institute of Health, Charité - Universitätsmedizin Berlin, Berlin, Berlin, Germany | 0000-0003-1311-5475 | Nothing to disclose. |
| Martin Holtkamp, MD *SeLECT Consortium* | Epilepsy-Center Berlin-Brandenburg, Department of Neurology with Experimental Neurology, Charité – Universitätsmedizin Berlin, Berlin, Germany. | 0000-0003-2258-1670 | Nothing reported. |
| Chiara Tatillo, MD *SeLECT Consortium* | Department of Neurology, Hôpital Universitaire de Bruxelles – Hôpital Erasme and Université Libre de Bruxelles, Brussels, Belgium | | Nothing to disclose. |
| Gilles Naeije, MD,PhD *SeLECT Consortium* | Neurology department, HUB-Hopital Erasme, Brussels, Belgium | 0000-0003-1580-1970 | Nothing to disclose. |
| Katarina Jood, MD PhD *International CVT Consortium* | Department of Neurology, Sahlgrenska University Hospital, Region Västra Götaland, Gothenburg. Sweden and Department of Clinical Neuroscience, Institute of Neuroscience and Physiology, Sahlgrenska Academy, University of Gothenburg, Gothenburg, Sweden. | 0000-0001-8746-1771 | KJ received funding from the Swedish state under the agreement between the Swedish government and the county councils (ALF agreement, ALFGBG-1006525) and grants from the Swedish Stroke Association, is a national principal investigator for the LIBREXIA STROKE study (Janssen), and received consultancy fees from Quantify. |
| Julian Conrad, MD *SeLECT Consortium* | Department of Neurology, University of Muenster, Muenster, Germany.; Division for Neurodegenerative Diseases, Department of Neurology, Universitätsmedizin Mannheim, University of Heidelberg, Mannheim, Germany. | 0000-0001-7912-0571 | Nothing reported. |
| Stefan Evers, MD PhD *SeLECT Consortium* | Department of Neurology, University of Muenster, Muenster, Germany Department of Neurology, Krankenhaus Lindenbrunn, Coppenbrügge, Germany | 0000-0003-4460-3244 | Nothing to disclose. |
| Andreas R. Luft, MD *SeLECT Consortium* | cereneo Center for Neurology and Rehabilitation, Vitznau, Switzerland Department of Health, Lake Lucerne Institute, Vitznau, Switzerland University Hospital of Zurich, Zurich, Switzerland | 0000-0001-9865-7382 | Nothing to disclose. |

| Name and network | Affiliation | ORCID | Disclosures |
|---|---|---|---|
| Miranda Stattmann, MD *SeLECT Consortium* | Department of Neurology, Clinical Neuroscience Center, University Hospital and University of Zurich, Zurich, Switzerland. | 0009-0007-4121-774X | Nothing reported. |
| Anton Schmick, MD FEBN *SeLECT Consortium* | Department of Neurology, Clinical Neuroscience Center, University Hospital and University of Zurich, Zurich, Switzerland. | 0000-0003-4087-2121 | Nothing reported. |
| Maria Pagiantza, MD *SeLECT Consortium* | Department of Neurology, Clinical Neuroscience Center, University Hospital and University of Zurich, Zurich, Switzerland. | | Nothing reported. |
| Giulio Bicciato, MD *SeLECT Consortium* | Department of Neurology, Clinical Neuroscience Center, University Hospital and University of Zurich, Zurich, Switzerland. | 0000-0002-2956-0189 | Nothing reported. |
| Lucia Sinka, MD *SeLECT Consortium* | Department of Neurology, Clinical Neuroscience Center, University Hospital and University of Zurich, Zurich, Switzerland. | 0000-0002-3724-3861 | Nothing reported. |
| Adam Strzelczyk, MD *SeLECT Consortium* | Epilepsy Center Frankfurt Rhine-Main, Department of Neurology, Goethe-University Frankfurt, Frankfurt am Main, Germany. | 0000-0001-6288-9915 | Nothing reported. |
| Carolina Ferreira-Atuesta, MD MSc *SeLECT Consortium* | Department of Neurology, Clinical Neuroscience Center, University Hospital Zurich and University of Zurich, Zurich, Switzerland | 0000-0003-0853-7163 | Nothing to disclose. |
| Mira Katan, MD, MSC *SeLECT Consortium* | University and University Hospital of Basel, Switzerland | 0000-0002-9265-8066 | Nothing reported. |
| Anna Cavallini, MD *MUCH-Italy Investigators* | Dept Emergency Neurology and Stroke Unit, IRCCS C. Mondino Foundation, Pavia, Italy. | 0000-0002-5227-1502 | Nothing to disclose. |
| Arianna Scartezzini, MD *SeLECT Consortium* | Stroke Unit, DAI di Neuroscienze, Azienda Ospedaliera Universitaria Integrata Verona, Verona, Italy. | 0009-0004-8492-1583 | Nothing to disclose. |
| Cecil D Hahn, MD, MPH *ILAE Acute Symptomatic Seizures Task Force* | Division of Neurology, The Hospital for Sick Children and Department of Paediatrics, University of Toronto. | 0000-0002-0887-8761 | Nothing to disclose. |
| Clio Aracelli Rubinos *ILAE Acute Symptomatic Seizures Task Force* | Department of Neurology, The University of North Carolina at Chapel Hill School of Medicine, Chapel Hill, NC, USA. | | Nothing to disclose. |
| Cristiano Azzini, MD *MUCH-Italy Investigators* | U.O. Neurologia, Stroke Unit, Azienda Ospedaliera Universitaria S. Anna Ferrara, Italia. | 0000-0003-2215-8143 | Nothing to disclose. |
| Dong Zhou *ILAE Acute Symptomatic Seizures Task Force* | Department of Neurology, West China Hospital, Sichuan University, Chengdu, Sichuan, China. | | Nothing to disclose. |
| Elisa Giorli, MD *MUCH-Italy Investigators* | Neurology Institution Sant’Andrea Hospital La Spezia. | | Nothing to disclose. |
| Federico Mazzacane, MD *SeLECT Consortium* | Dept Emergency Neurology and Stroke Unit, IRCCS C. Mondino Foundation, Pavia, Italy. | 0000-0002-2224-8002 | Nothing to disclose. |

| **Name and network** | **Affiliation** | **ORCID** | **Disclosures** |
|---|---|---|---|
| Francesco Valletta, MD *SeLECT Consortium* | Stroke Unit, DAI di Neuroscienze, Azienda Ospedaliera Universitaria Integrata Verona, Verona, Italy. | 0009-0005-1568-9669 | Nothing to disclose. |
| Gagandeep Singh, DM *ILAE Acute Symptomatic Seizures Task Force* | Department of Neurology, Dayanand Medical College & Hospital, Ludhiana, India. | 0000-0001-6661-3553 | Nothing to disclose. |
| Giacomo Giacalone, MD, PhD *MUCH-Italy Investigators* | Stroke Unit, U.O. Neurologia, IRCCS Ospedale San Raffaele, Milano, Italy. | | Nothing to disclose. |
| Giorgio Silvestrelli, MD PhD *MUCH-Italy Investigators* | Neurology Unit, Emergency Department, Imperia Hospital, ATS Liguria, Italy. | 0000-0003-2060-6416 | Nothing to disclose. |
| Hui Jan Tan, MD, FRCP *SeLECT Consortium* | Department of Medicine, Faculty of Medicine, Universiti Kebangsaan Malaysia (The National University of Malaysia), and Department of Medicine, Hospital Canselor Tuanku Muhriz, Kuala Lumpur, Malaysia. | 0000-0003-4827-7984 | Nothing to disclose. |
| Jo Wilmshurst, MB BS, MRCPaed, FCPaed, MD *ILAE Acute Symptomatic Seizures Task Force* | Department of Paediatric Neurology, Red Cross War Memorial Children's Hospital, Neuroscience Institute, University of Cape Town, South Africa. | 0000-0001-7328-1796 | Nothing to disclose. |
| Johan Zelano, MD PhD *ILAE Acute Symptomatic Seizures Task Force* | 1. Department of Clinical Neuroscience, Institute of Neuroscience and Physiology, the; Sahlgrenska Academy at the University of Gothenburg, Gothenburg, Sweden; 2. Department of Neurology, Sahlgrenska University Hospital, member of ERN Epicare,; Gothenburg, Sweden; 3. Wallenberg Center of Molecular and Translational Medicine, Gothenburg University,; Sweden. | 0000-0001-9445-4545 | JZ has received advisory board and speaker honoraria from UCB, Eisai, Orion Pharma, Angelini Pharma, and Sanofi for non-branded education; as an employee of Sahlgrenska University Hospital, he is or has been an investigator or sub-investigator in clinical trials sponsored by GW Pharma, SK Life Science, UCB, Angelini Pharma, and Bial (no personal compensation). |

| Name and network | Affiliation | ORCID | Disclosures |
|---|---|---|---|
| Lawrence J. Hirsch, MD *ILAE Acute Symptomatic Seizures Task Force* | Comprehensive Epilepsy Center, Department of Neurology, Yale University School of Medicine, New Haven, CT, USA. | 0000-0002-6333-832X | LJH reports research support to Yale University for investigator-initiated studies from the Yale NORSE/FIRES Biorepository Fund, the National Institutes of Health, and the Patient-Centered Outcomes Research Institute; consultation fees from Ceribell, Natus, Tisento, and UCB; royalties from Wolters Kluwer (UpToDate), Wiley (Atlas of EEG in Critical Care), and Springer (associate editor, Epilepsy Surgery: A Practical Case-based Approach); and honoraria for speaking or training from NeuroPace and Natus. LJH is co-chair of the medical and scientific advisory board of the NORSE Institute. |
| Luis Otavio Caboclo *ILAE Acute Symptomatic Seizures Task Force* | Hospital Israelita Albert Einstein, São Paulo, SP, Brazil. | | Nothing to disclose. |
| Manuel Cappellari, MD *MUCH-Italy Investigators* | Stroke Unit, DAI di Neuroscienze, Azienda Ospedaliera Universitaria Integrata, Piazzale A. Stefani 1, 37126 Verona, Italy. | 0000-0002-3534-3201 | Nothing to disclose. |
| Marialuisa Zedde, MD *MUCH-Italy Investigators* | Neurology Unit, Stroke Unit, AUSL-IRCCS di Reggio Emilia, Reggio Emilia, Italy; Neurology Unit and Stroke Unit, Ospedale del Mare, ASL Napoli 1 Centro, Napoli, Italy. | 0000-0001-7530-818X | Nothing to disclose. |
| Markus Arnold, MD *SeLECT Consortium* | Department of Neurology and Stroke Center, University Hospital of Basel and University of Basel, Basel, Switzerland. | | Nothing to disclose. |
| Massimo Del Sette, MD *MUCH-Italy Investigators* | AOM IRCCS Ospedale Policlinico San Martino. | | Nothing to disclose. |
| Maurizio Paciaroni, MD *MUCH-Italy Investigators* | Department of Neurosciences and Rehabilitation, University of Ferrara, Ferrara - Italy. | 0000-0002-5483-8795 | Nothing to disclose. |
| Mauro Magoni, MD *MUCH-Italy Investigators* | Stroke Unit, Neurologia Vascolare, Spedali Civili di Brescia, Italia | | Nothing to disclose. |
| Melody T Asukile, MD *ILAE Acute Symptomatic Seizures Task Force* | Department of Internal Medicine, University of Zambia School of Medicine, Lusaka, Zambia; UCL Queen Square Institute of Neurology, London, UK. | 0000-0003-3745-8982 | Nothing to disclose. |
| Michael Knoflach, MD *SeLECT Consortium* | Department of Neurology, Medical University of Innsbruck, Innsbruck, Austria; and VASCage, Centre on Clinical Stroke Research, Innsbruck, Austria. | 0000-0001-5576-6562 | Nothing to disclose. |

| Name and network | Affiliation | ORCID | Disclosures |
|---|---|---|---|
| Nick Abend *ILAE Acute Symptomatic Seizures Task Force* | Departments of Neurology, Anesthesia and Critical Care Medicine, and Pediatrics, Children's Hospital of Philadelphia, Philadelphia, PA, USA. | | Nothing to disclose. |
| Nicola Specchio *ILAE Acute Symptomatic Seizures Task Force* | Department of Neuroscience, Bambino Gesù Children's Hospital, IRCCS, Full Member of the European Reference Network on Rare and Complex Epilepsies (EpiCARE), Rome, Italy. | | Nothing to disclose. |
| Nirmeen A. Kishk *ILAE Acute Symptomatic Seizures Task Force* | Neurology Department, Faculty of Medicine, Cairo University, Cairo, Egypt. | | Nothing to disclose. |
| Patrick Kwan, MD PhD *SeLECT Consortium* | Department of Neuroscience, Monash Institute of Medical Engineering, Monash University, Melbourne, Australia. | | Nothing to disclose. |
| Rocco Salvatore Calabrò, MD, PhD *MUCH-Italy Investigators* | Istituto di Ricovero e Cura a Carattere Scientifico, Centro Neurolesi Bonino-Pulejo, Messina, Italia | | Nothing to disclose. |
| Rossana Tassi, MD *MUCH-Italy Investigators* | Stroke Unit, Department of Emergency Urgency, AOU Senese, Siena Italy. | 0000-0002-5906-8718 | RT reports consultancy fees from Boehringer Ingelheim, AstraZeneca, and Daiichi-Sankyo. |
| Ruta Yardi *ILAE Acute Symptomatic Seizures Task Force* | Department of Neurology, Geisinger Neuroscience Institute, Danville, PA, USA. | | Nothing to disclose. |
| Sahar Zafar *ILAE Acute Symptomatic Seizures Task Force* | Value Based Health Care Center, Neurology Comparative Effectiveness Research Lab, Department of Neurology, Massachusetts General Hospital and Harvard Medical School, Boston, MA, USA. | | Nothing to disclose. |
| Simona Lattanzi, MD PhD *ILAE Acute Symptomatic Seizures Task Force* | Neurological Clinic, Department of Experimental and Clinical Medicine, Marche Polytechnic University, Ancona, Italy. | 0000-0001-8748-0083 | Nothing to disclose. |
| Simona Marcheselli, MD *MUCH-Italy Investigators* | Neurologia d'urgenza e Stroke UNit, IRCCS Humanitas Research Hospital, Rozzano, MI, Italy. | 0000-0002-7238-4088 | Nothing to disclose. |
| Simone Vidale, MD *MUCH-Italy Investigators* | Department of Neurology & Stroke Unit, ASST Sette Laghi, Varese, Italy. | 0000-0003-1426-0885 | Nothing to disclose. |
| Stephane Auvin *ILAE Acute Symptomatic Seizures Task Force* | Assistance Publique-Hôpitaux de Paris, Hôpital Universitaire Robert-Debré, Paris, France; Institut Hospitalo-Universitaire Robert-Debré du Cerveau de l'Enfant, Paris, France; Université Paris Cité, INSERM NeuroDiderot, Paris, France; Institut Universitaire de France, Paris, France. | | Nothing to disclose. |
| Susanne Knake, MD *ILAE Acute Symptomatic Seizures Task Force* | Philipps Universität Marburg, Department of Neurology, Epilepsy Center Hessen, Marburg, Germany. | 0000-0002-6298-0513 | SK received speaker honoraria from Angelini Pharma, Bial, Eisai, Jazz Pharma, Kanso, Merck Serono, and UCB. |
| Yamile Calle-Lopez, MD *ILAE Acute Symptomatic Seizures Task Force* | Facultad de Medicina, Universidad de Antioquia UdeA, Calle 70 No. 52-21, Medellín, Colombia. Hospital San Vicente Fundacion, Medellín, Colombia. | 0000-0002-7805-9200 | Nothing to disclose. |
| Zhen Li, MMed *SeLECT Consortium* | Department of Neurology, Nanjing Drum Tower Hospital, Affiliated Hospital of Medical School, Nanjing University, Nanjing, China. | | Nothing to disclose. |

| Name and network | Affiliation | ORCID | Disclosures |
|---|---|---|---|
| Turgut Tatlisumak, MD PhD *International CVT Consortium* | 1. Department of Clinical Neuroscience, Institute of Neuroscience and Physiology, Sahlgrenska Academy at University of Gothenburg and Department of Neurology, Sahlgrenska University Hospital, Gothenburg, Sweden. 2. Department of Neurology, Helsinki University Hospital and University of Helsinki, Helsinki, Finland. | | Nothing to disclose. |
| Marcel Arnold, MD PhD *International CVT Consortium* | Department of Neurology, Inselspital, Bern University Hospital and University of Bern, Bern, Switzerland | | Nothing to disclose. |
| José M Ferro, MD PhD *International CVT Consortium* | Instituto de Medicina Molecular João Lobo Antunes, Faculdade de Medicina, Universidade de Lisboa, Portugal | | Nothing to disclose. |
| Patrícia Canhão, MD PhD *International CVT Consortium* | Faculdade de Medicina, Universidade de Lisboa, Lisbon, Portugal | | Nothing to disclose. |
| Antonio Arauz, MD PhD *International CVT Consortium* | National Institute of Neurology and Neurosurgery Manuel Velasco Suarez, Mexico-City, Mexico | | Nothing to disclose. |
| Masoud Ghiasian, MD *International CVT Consortium* | Sina Hospital, Hamadan University of Medical Sciences, Hamadan, Iran | | Nothing to disclose. |
| Maryam Mansour, MD *International CVT Consortium* | Sina Hospital, Hamadan University of Medical Sciences, Hamadan, Iran | | Nothing to disclose. |
| Daniel Guisado-Alonso, MD *International CVT Consortium* | Department of Neurology, Hospital de la Santa Creu i Sant Pau, Barcelona, Spain | | Nothing to disclose. |
| Esme Ekizoglu, MD *International CVT Consortium* | Department of Neurology, Istanbul Faculty of Medicine, Istanbul University, Istanbul, Türkiye | | Nothing to disclose. |
| Nilüfer Yesilot, MD *International CVT Consortium* | Department of Neurology, Istanbul Faculty of Medicine, Istanbul University, Istanbul, Türkiye | | Nothing to disclose. |
| Mine Sezgin, MD *International CVT Consortium* | Department of Neurology, Istanbul Faculty of Medicine, Istanbul University, Istanbul, Türkiye | | Nothing to disclose. |
| Miguel A Barboza, MD MSc *International CVT Consortium* | Neurosciences Department, Hospital Dr. R.A. Calderón Guardia, CCSS, San José, Costa Rica | | Nothing to disclose. |
| Valentina Arnao, MD PhD *International CVT Consortium* | Department of Biomedicine, Neuroscience and Advanced Diagnostics, University of Palermo, Palermo, Italy | | Nothing to disclose. |
| Paolo Aridon, MD PhD *International CVT Consortium* | Department of Biomedicine, Neuroscience and Advanced Diagnostics, University of Palermo, Palermo, Italy | | Nothing to disclose. |
| Carlos Garcia-Esperon, MD *International CVT Consortium* | Department of Neurology, Priority Research Centre for Brain and Mental Health Research, John Hunter Hospital, University of Newcastle, Newcastle, NSW, Australia | | Nothing to disclose. |
| João Morgadinho, MD *International CVT Consortium* | Department of Neurology, Hospital Garcia de Orta, Local Health Unit, Almada-Seixal, Portugal | 0009-0004-4837-819X | Nothing to disclose. |
| Miguel Rodrigues, MD *International CVT Consortium* | Department of Neurology, Hospital Garcia de Orta, Local Health Unit, Almada-Seixal, Portugal | | Nothing to disclose. |
